# Examining the association of live virus neutralisation activity of capillary microsamples and risk of SARS-CoV-2 infections: a nested case control study within the Virus Watch community cohort

**DOI:** 10.1101/2023.11.28.23299156

**Authors:** Alexei Yavlinsky, Vincent G. Nguyen, Sarah Beale, Emma Wall, Mary Y Wu, Isobel Braithwaite, Jana Kovar, Madhumita Shrotri, Annalan M D Navaratnam, Wing Lam Erica Fong, Thomas E. Byrne, François Balloux, Ibrahim Abubakar, Benjamin J. Cowling, Andrew C. Hayward, Robert W. Aldridge

**Author notes:** Corresponding Author: Robert W. Aldridge, Centre for Public Health Data Science, Institute of Health Informatics, University College London, NW1 2DA, UK.

## Abstract

Due to the proliferation of new SARS-CoV-2 variants, most COVID-19 cases are now caused by post-vaccine infections and a substantial proportion are reinfections. While prior research on correlates of protection has focused on the role of anti-spike antibodies, the results of the corresponding antibody assays may not accurately predict the risk of infection with new SARS-CoV-2 variants. In this study, we investigated the association between live virus neutralising antibody activity and SARS-CoV-2 infection risk using self-administered capillary microsample blood tests from VirusWatch participants. The study was conducted during the transition between the dominance of the B.1.617.2 (Delta) and B.1.1.529 (Omicron BA.1) SARS-CoV-2 variants, enabling us to investigate the association between variant-specific virus inhibition and subsequent infections within each dominance period. Greater inhibition of Omicron BA.1 live virus was associated with a reduction in infection risk during both the Delta and Omicron BA.1 dominance periods. Delta virus inhibition was associated with infection risk reduction during the Delta dominance period, but we found no association between Delta inhibition and protection against infection during the Omicron BA.1 dominance period. Our results are consistent with earlier findings and suggest that variant-specific serosurveillance of immunity and protection against SARS-CoV-2 infection at the population level could inform public health policy in near-real time using inexpensive and accessible home-based testing.

## Introduction

Due to high population vaccination rates in the UK (up to 88% vaccinated with two doses), the majority of COVID-19 cases are now likely caused by post-vaccine infections (1). In England and Wales in October 2023, over 41% of COVID-19 cases were reinfections (2). This is largely attributable to the proliferation of new SARS-CoV-2 variants (3–6). Previous work on defining possible correlates of protection against SARS-CoV-2 infection has focused on recency of vaccination and spike binding (anti-spike) antibody levels (7) and on neutralising antibody titres following primary infection and Alpha reinfection (8). However, variant-related changes in infectivity and evolving viral immune escape require continuous re-evaluation of antibody mediated protection (9).

Increased anti-spike antibody levels have been shown to be associated with a reduced risk of acquiring a SARS-CoV-2 infection (7,10). However, the assays for measuring anti-S binding antibodies were developed using the ancestral strain of the SARS-CoV-2 virus, with titres therefore having limited association with the risk of being infected with new variants such as Omicron BA.1 (11). Neutralising antibody titres to specific variants may offer a more precise and up-to-date measure of the degree of protection against infection with these strains (12), as these antibodies are likely to play a major role in protecting against infection, are associated with survival from COVID-19 and are the most widely recognised working correlate of protection (12,13). While research on SARS-CoV-2 correlates of protection is ongoing, there is an urgent need to better define populations who continue to be at risk of severe COVID-19 for targeted vaccination policies (14).

In this paper, we used VirusWatch study participants’ self-administered capillary microsample blood tests to examine the association between live virus neutralising antibody activity of these microsamples and participants’ risk of SARS-CoV-2 infection. Notably, data collection was performed during the transition between the dominance of the B.1.617.2 (Delta) and B.1.1.529 (Omicron BA.1) SARS-CoV-2 variants, enabling us to investigate the association between variant-specific virus inhibition and subsequent infections within each period, with a view to developing home-based screening methods for at-risk populations for targeted vaccination and informing public health policy.

## Methods

### Study design and setting

The Virus Watch study adheres to all relevant ethical regulations and the study protocol has been approved by the Hampstead NHS Health Research Authority Ethics Committee (ethics approval number—20/HRA/2320). Informed consent was obtained from participants. This study is a nested case control study within Virus Watch, a prospective household community cohort study investigating acute respiratory infections in England & Wales that started recruitment in June 2020. A detailed description of recruitment methods has been described previously (15), but in summary, to recruit participants we used a range of methods. We used the Royal Mail Post Office Address File to generate a random list of residential address lists that were sent recruitment postcards, we placed social media adverts on Facebook and Twitter and sent SMS messages and letters to participants from their General Practitioners. Recruitment ended in March 2022 and active follow up is ongoing.

### Data sources

Participants in this study were followed-up weekly by email with a link to an illness survey which asked about the presence or absence of symptoms that could indicate COVID-19 infection.

Symptoms were grouped by respiratory, gastrointestinal and general infection symptoms. SARS-CoV-2 test results received from outside the study were also captured via linking survey data with the Second Generation Surveillance System, the national testing database for England and Wales. Vaccination data was obtained from self-reporting via the weekly survey and from the linkage to the National Immunisation Management System. Where available, linkage results were preferentially chosen over self-reporting. Further details of linkage methods are specified in (16).

### Participants

Nested within the larger Virus Watch study is a sub-cohort of 19,457 adults (aged 18 years or over) participating in antibody testing who completed and returned at least one at-home capillary blood sampling kit (Thriva Ltd.). Kits were sent to participants via post on a monthly basis from February-August 2021 and every other month thereafter until April 2022. These microsamples were originally analysed for the presence of anti-S and anti-N SARS-CoV-2 antibodies. For this study, we analysed a subset of these microsamples, taken between November 3, 2021 and March 11, 2022, within the context of a case-control study design.

### Case selection

Cases were selected on the basis of developing a SARS-CoV-2 infection, confirmed through a positive PCR or LFD test result between November 3, 2021 and March 11, 2022. Positive tests were identified either by participant self-report or from linkage of patient demographic characteristics (name, date of birth, address, NHS number) to the national Second Generation Surveillance System for SARS-CoV-2. For cases, the closest microsample prior to the date of the positive test was chosen.

### Control selection

Our initial strategy for choosing controls was to select those individuals who never received or reported a positive PCR or LFD test result throughout the duration of the study. We group-matched cases to controls in a 1:3 ratio, based on age, sex and whether they resided in or outside of London at study enrolment. For controls, the most recent available microsample was selected.

The strategy for sampling cases and controls was designed prior to the first detection of the Omicron BA.1 variant in the UK. To estimate variant-specific infection risk and address time-varying confounding, we further selected controls who tested negative within the corresponding variant dominance periods. Controls without a recorded negative PCR or LFD test result were therefore excluded.

### Study periods

For the scope of this study, we defined two dominance periods: Delta (November 3 - December 6, 2021) and Omicron BA.1 (December 15, 2021 - February 17, 2022). To define these periods, we used the UKHSA Delta to Omicron BA.1 dominance transition date of December 11, 2021 and the Omicron BA.1 to Omicron BA.2 transition date of February 22, 2022 (6) and added/subtracted a gap of four days to each date (total gap of eight days between Delta and Omicron BA.1) to avoid analysing samples when both variants were circulating simultaneously.

For each period, we selected the cases and controls who had completed both the capillary microsample tests and the PCR/LFD tests between the associated start and end dates, resulting in two separate subgroups to be analysed. There were an insufficient number of individuals submitting capillary microsamples and PCR/LFD test results to calculate infection risk during Omicron BA.2 variant dominance, commencing February 22, 2022.

### Exposures

Age and sex were taken from participants’ responses to demographic questions at study baseline. Vaccination status was derived from either participant self-report or from linkage of patient demographic characteristics (name, date of birth, address, NHS number) to the National Immunisation Management Service (NIMS). We categorised people as clinically vulnerable (CV) or clinically extremely vulnerable (CEV) using our previously reported methods (7). We used official criteria specified by the UK Government to categorise people based on their level of vulnerability to COVID-19. These criteria used guidelines from Public Health England (now the United Kingdom Health Security Agency), and the Department of Health and Social Care to identify those who were clinically extremely vulnerable (17), and guidelines from the Joint Committee on Vaccination and Immunisation to identify those who were clinically vulnerable (18). The criteria were adapted based on clinical variables collected using baseline and monthly surveys. Individuals who did not meet the criteria for being clinically extremely vulnerable were placed in the clinically vulnerable group, and those who did not meet either criteria were considered to have no sign of clinical vulnerability. Individuals for whom there was not enough data on clinical characteristics were included in the not clinically vulnerable group.

### Sample testing

Microsamples were tested using a high-throughput live SARS-CoV-2 neutralisation assay to determine neutralising antibody titres (NAbTs) against Delta, Omicron BA.1 and BA.2 variants. For each of the microsamples, median 50% inhibitory concentration (IC50) for the three variants was obtained (19); these were the primary exposure variables.

### Statistical methods

Individuals whose microsamples failed on the live virus assay for a specific variant were excluded from the subsequent analyses for that variant. Microsamples that had weak or no detectable inhibitory activity were assigned the IC50 value of 30 and those that had shown complete inhibition were given the IC50 value of 3000, reflecting the quantitative range of the assays (40–2560). The IC50 values were log-transformed in base two. To examine the properties of the live virus tests and inform our analyses, we examined the distribution of log_2_(IC50) values by case/control status, live virus assay variant and dominant variant time period. Logistic regression was used for crude and multivariable analyses. For the latter, the models were adjusted for age, sex and clinical vulnerability status confounders. We also adjusted for whether the individual received a booster prior to reporting a PCR/LFD test result, which in this case can be considered an effect modifier.

For each subgroup, we calculated the mean log_2_(IC50) for each variant, stratified by covariate categories and whether the individual had received a booster prior to taking their microsample test.

For each live virus assay variant, we calculated protection curves within each dominance period, using infection odds ratios for log_2_(IC50) values that were stratified into quintiles. These odds ratios were adjusted age, sex and clinical vulnerability.

For each subgroup, we performed a sensitivity analysis to characterise the effect of gap duration between dominance periods on our multivariable model estimates.

For each subgroup and live virus assay variant, we examined how log_2_(IC50) values changed over time after receiving a booster dose and before testing positive for SARS-CoV-2.

## Results

Capillary microsamples were received from 13021 individuals, who used test kits sent out between November 1st, 2021 and February 28, 2022, with tests taken on dates ranging from November 3, 2021 - March 5, 2022 (Figure 1). 24284 capillary microsample tests were returned in this period. Within this sample, we identified 1243 cases of SARS-CoV-2 and matched 3729 controls. We were able to locate 1216 cases and 3654 control samples in storage, and deemed 1015 cases and 3062 controls as having sufficient volume for further analysis. Upon sample receipt at the Francis Crick Institute, 91 cases and 208 controls had insufficient volume for assay processing. We were therefore able to analyse samples for 924 cases and 2854 controls. 100 cases and 304 controls failed the Delta live virus assay, 166 cases and 482 controls failed the Omicron BA.1 assay and 186 cases and 520 controls failed the Omicron BA.2 assay.

**Figure 1.**
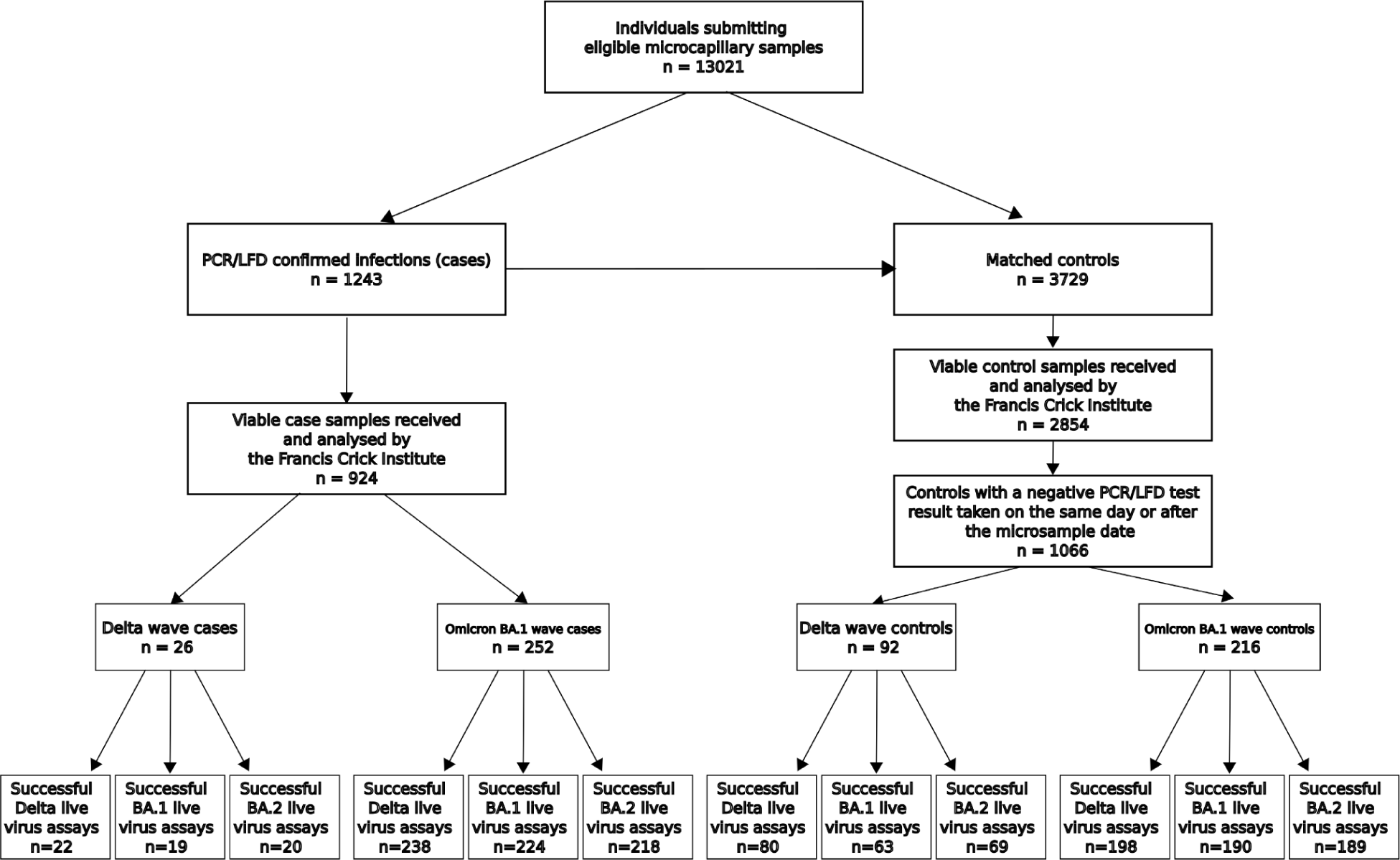
Flow diagram representing participants’ selection into the study.

Table 1 presents the demographic and vaccination characteristics for eligible, selected and analysed participants, and Table 2 presents the same characteristics for the selected and analysed participants split by case/control status. Stratum proportions were uniform for all exposures when comparing selected and analysed participants, and this relationship was maintained when the participants were split by case/control status. This indicates that discarding microsamples with insufficient volume likely did not introduce selection bias for the exposure variables. Amongst analysed participants (N = 3778), there were more women than men (59% vs 41%) and the largest age group was 45-64 years (44%), followed by 65+ years (36%). Most analysed participants were white British (90%) and most had ChAdOx1 (AstraZeneca) as their second vaccine dose (55%). Most participants had a third mRNA vaccine dose before or during the study (81%).

**Table 1.** Demographic and vaccination characteristics of eligible and analysed participants.

|  | **Eligible** | **Selected** | **Analysed** |
| --- | --- | --- | --- |
| **Exposure** | N = 13,021<sup>1</sup> | N = 4,972<sup>1</sup> | N = 3,778<sup>1</sup> |
| **Age group** |  |  |  |
| 16-24 | 165 (1.3%) | 80 (1.6%) | 67 (1.8%) |
| 25-44 | 1,516 (12%) | 794 (16%) | 636 (17%) |
| 45-64 | 5,620 (43%) | 2,219 (45%) | 1,708 (45%) |
| 65+ | 5,720 (44%) | 1,879 (38%) | 1,367 (36%) |
| **Sex** |  |  |  |
| Female | 7,507 (58%) | 2,932 (59%) | 2,243 (59%) |
| Intersex | 6 (<0.1%) |  |  |
| Male | 5,138 (39%) | 2,040 (41%) | 1,535 (41%) |
| Missing | 365 (2.8%) |  |  |
| Prefer not to say | 5 (<0.1%) |  |  |
| **Ethnicity** |  |  |  |
| Black | 37 (0.3%) | 16 (0.3%) | 12 (0.3%) |
| Missing | 367 (2.8%) | 2 (<0.1%) |  |
| Mixed | 102 (0.8%) | 53 (1.1%) | 46 (1.2%) |
| Other Asian | 57 (0.4%) | 27 (0.5%) | 19 (0.5%) |
| Other Ethnicity | 33 (0.3%) | 18 (0.4%) | 14 (0.4%) |
| Prefer not to say | 31 (0.2%) | 15 (0.3%) | 10 (0.3%) |
| South Asian | 90 (0.7%) | 47 (0.9%) | 37 (1.0%) |
| White British | 11,613 (89%) | 4,457 (90%) | 3,388 (90%) |
| White Irish | 147 (1.1%) | 62 (1.2%) | 40 (1.1%) |
| White Other | 544 (4.2%) | 275 (5.5%) | 212 (5.6%) |
| **Clinical vulnerability** |  |  |  |
| Clinically extremely vulnerable | 1,360 (10%) | 503 (10%) | 372 (9.8%) |
| Clinically vulnerable | 3,706 (28%) | 1,422 (29%) | 1,061 (28%) |
| Missing | 21 (0.2%) | 6 (0.1%) | 4 (0.1%) |
| Not clinically vulnerable | 7,934 (61%) | 3,041 (61%) | 2,341 (62%) |
| **Location** |  |  |  |
| London | 1,214 (9.3%) | 620 (12%) | 474 (13%) |
| **Exposure** | N = 13,021<sup>1</sup> | N = 4,972<sup>1</sup> | N = 3,778<sup>1</sup> |
| Not London | 11,807 (91%) | 4,352 (88%) | 3,304 (87%) |
| **2nd dose vaccine type** |  |  |  |
| BNT162b2 | 4,041 (31%) | 1,563 (31%) | 1,208 (32%) |
| ChadOx1 | 6,924 (53%) | 2,758 (55%) | 2,071 (55%) |
| Don't know / don't remember | 23 (0.2%) | 13 (0.3%) | 10 (0.3%) |
| Missing | 1,886 (14%) | 569 (11%) | 438 (12%) |
| mRNA-127 | 84 (0.6%) | 40 (0.8%) | 31 (0.8%) |
| Other | 63 (0.5%) | 29 (0.6%) | 20 (0.5%) |
| **Had booster before/during study** | 10,234 (79%) | 4,023 (81%) | 3,045 (81%) |

<sup>1</sup>n (%)

**Table 2.** Demographic and vaccination characteristics of the selected and analysed participants, split by case/control status.

| Exposure | Selected |  | Analysed |  |
| --- | --- | --- | --- | --- |
|  | Case, N = 1,243<sup>1</sup> | Control, N = 3,729<sup>1</sup> | Case, N = 924<sup>1</sup> | Control, N = 2,854<sup>1</sup> |
| **Age group** |  |  |  |  |
| 16-24 | 21 (1.7%) | 59 (1.6%) | 17 (1.8%) | 50 (1.8%) |
| 25-44 | 216 (17%) | 578 (16%) | 166 (18%) | 470 (16%) |
| 45-64 | 552 (44%) | 1,667 (45%) | 420 (45%) | 1,288 (45%) |
| 65+ | 454 (37%) | 1,425 (38%) | 321 (35%) | 1,046 (37%) |
| **Sex** |  |  |  |  |
| Female | 733 (59%) | 2,199 (59%) | 539 (58%) | 1,704 (60%) |
| Male | 510 (41%) | 1,530 (41%) | 385 (42%) | 1,150 (40%) |
| **Ethnicity** |  |  |  |  |
| Black, Other Asian, Other Ethnicity | 19 (1.5%) | 42 (1.1%) | 14 (1.5%) | 31 (1.1%) |
| Missing | 1 (<0.1%) | 1 (<0.1%) |  |  |
| Mixed | 13 (1.0%) | 40 (1.1%) | 11 (1.2%) | 35 (1.2%) |
| Prefer not to say | 1 (<0.1%) | 14 (0.4%) | 1 (0.1%) | 9 (0.3%) |
| South Asian | 16 (1.3%) | 31 (0.8%) | 12 (1.3%) | 25 (0.9%) |
| White British | 1,117 (90%) | 3,340 (90%) | 830 (90%) | 2,558 (90%) |
| White Irish | 16 (1.3%) | 46 (1.2%) | 9 (1.0%) | 31 (1.1%) |
| White Other | 60 (4.8%) | 215 (5.8%) | 47 (5.1%) | 165 (5.8%) |
| **Clinical vulnerability** |  |  |  |  |
| Clinically extremely vulnerable | 120 (9.7%) | 383 (10%) | 87 (9.4%) | 285 (10.0%) |
| Clinically vulnerable | 356 (29%) | 1,066 (29%) | 254 (27%) | 807 (28%) |
| Missing | 0 (0%) | 6 (0.2%) | 0 (0%) | 4 (0.1%) |
| Not clinically vulnerable | 767 (62%) | 2,274 (61%) | 583 (63%) | 1,758 (62%) |
| **Location** |  |  |  |  |
| London | 155 (12%) | 465 (12%) | 119 (13%) | 355 (12%) |
| Not London | 1,088 (88%) | 3,264 (88%) | 805 (87%) | 2,499 (88%) |
| **2nd dose vaccine type** |  |  |  |  |
| BNT162b2 | 406 (33%) | 1,157 (31%) | 305 (33%) | 903 (32%) |

| **Exposure** | **Selected** |  | **Analysed** |  |
| --- | --- | --- | --- | --- |
|  | Case, N = 1,243<sup>1</sup> | Control, N = 3,729<sup>1</sup> | Case, N = 924<sup>1</sup> | Control, N = 2,854<sup>1</sup> |
| ChadOx1 | 677 (54%) | 2,081 (56%) | 496 (54%) | 1,575 (55%) |
| Don't know / don't remember | 5 (0.4%) | 8 (0.2%) | 4 (0.4%) | 6 (0.2%) |
| Missing | 133 (11%) | 436 (12%) | 104 (11%) | 334 (12%) |
| mRNA-127 | 14 (1.1%) | 26 (0.7%) | 10 (1.1%) | 21 (0.7%) |
| Other | 8 (0.6%) | 21 (0.6%) | 5 (0.5%) | 15 (0.5%) |
| **Had booster before/during study** | 1,004 (81%) | 3,019 (81%) | 737 (80%) | 2,308 (81%) |

<sup>1</sup>n (%)

There were 2230 controls who had negative PCR or LFD test results during the analytical period of the study and 1066 had submitted their capillary microsamples prior to or on the same day as reporting negative PCR or LFD results. There were 26 cases and 92 controls for the Delta dominance period, with 22 cases and 80 controls analysed successfully on the Delta live virus assay, 19 cases and 63 controls analysed successfully on the Omicron BA.1 assay, and 20 cases and 69 controls analysed successfully on the Omicron BA.2 assay. There were 252 cases and 216 controls for the Omicron BA.1 period, with 238 cases and 198 controls analysed successfully on the Delta live virus assay, 224 cases and 190 controls analysed successfully on the Omicron BA.1 assay, and 218 cases and 189 controls analysed successfully on the Omicron BA.2 assay. Table 3 shows participants’ baseline characteristics for the two dominance periods and Table 4 splits these characteristics further by case/control status.

**Table 3.**
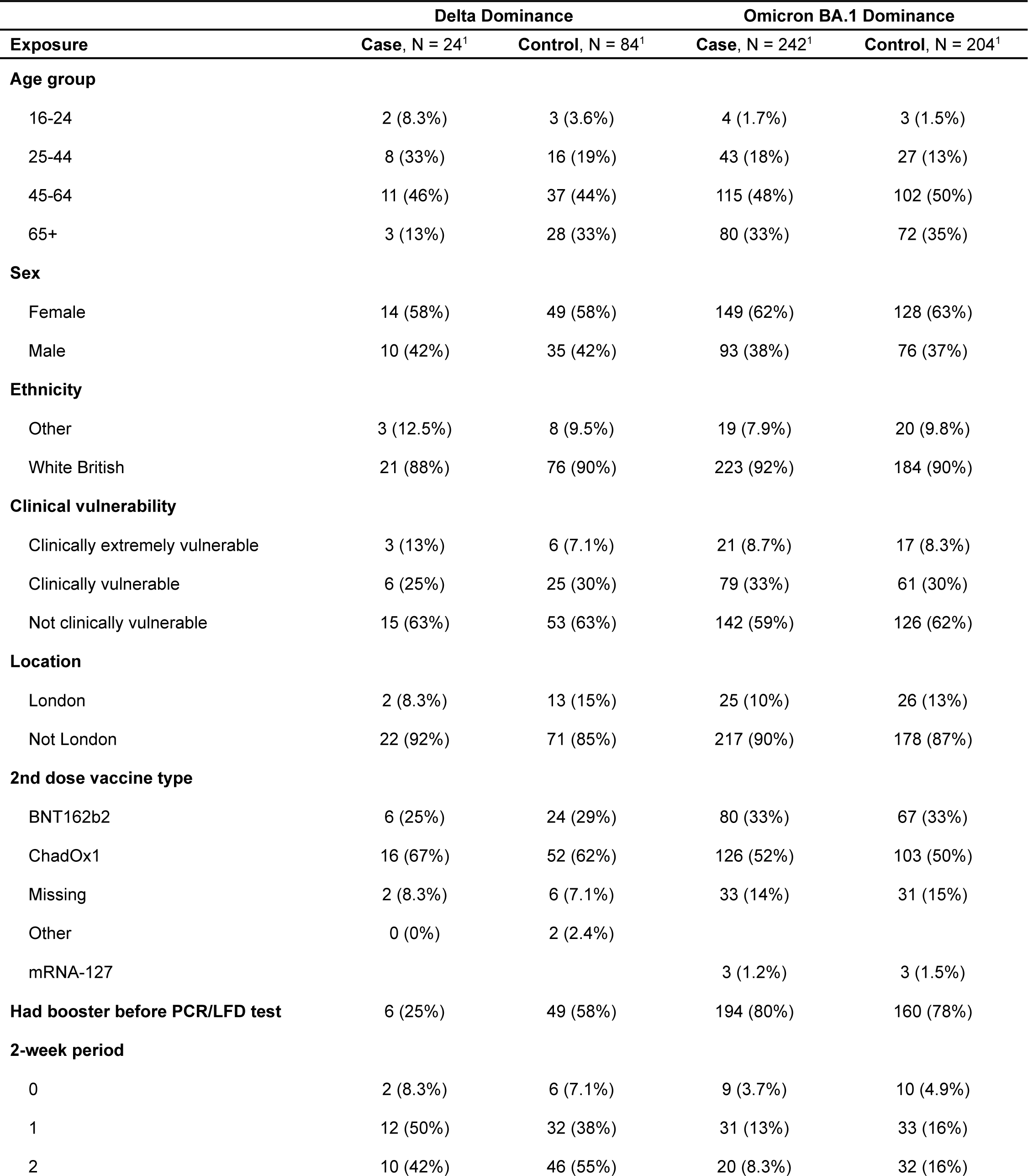
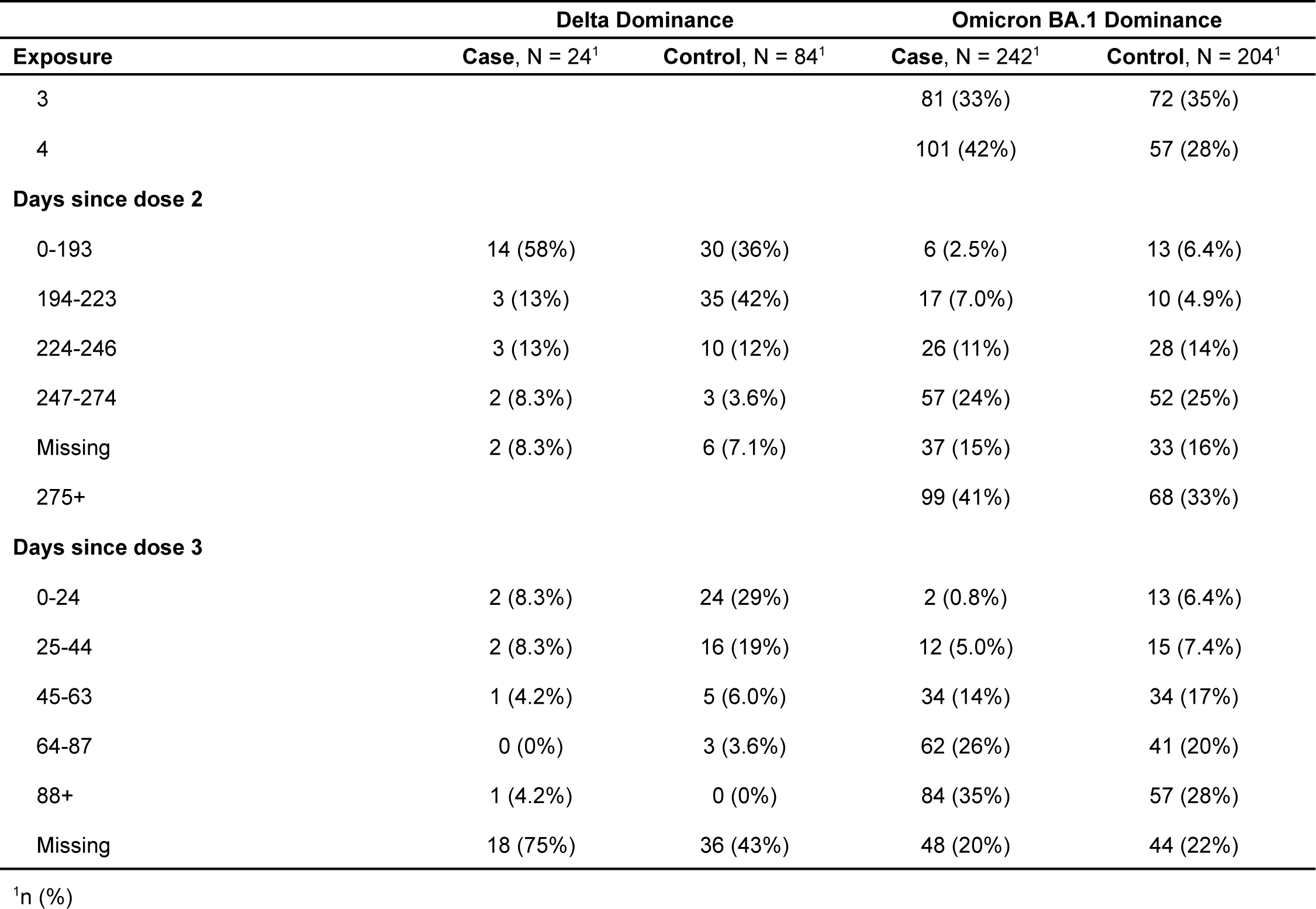
Participants’ baseline characteristics in the two subgroups submitting capillary microsamples during the Delta and Omicron BA.1 dominance periods. Participants whose samples were successful on at least one of the three live virus assays were included in this summary. The “2-week period” variable specifies the number of individuals with a recorded PCR/LFD test within given fortnightly segments of the corresponding dominance period. “Days since dose 2” and “days since dose 3” refer to the number of days elapsed between the vaccine dose and the PCR/LFD test.

**Table 4.**
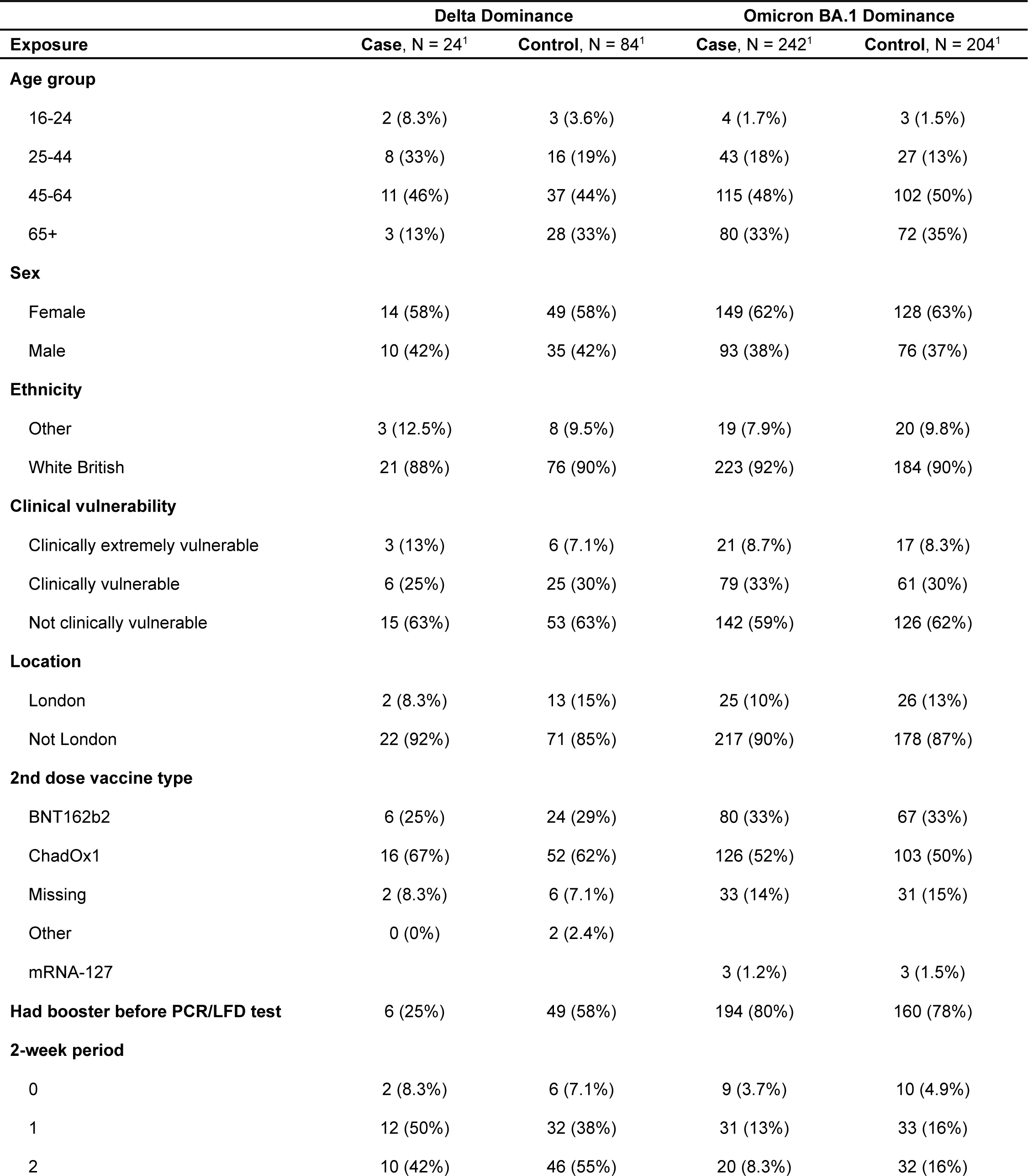
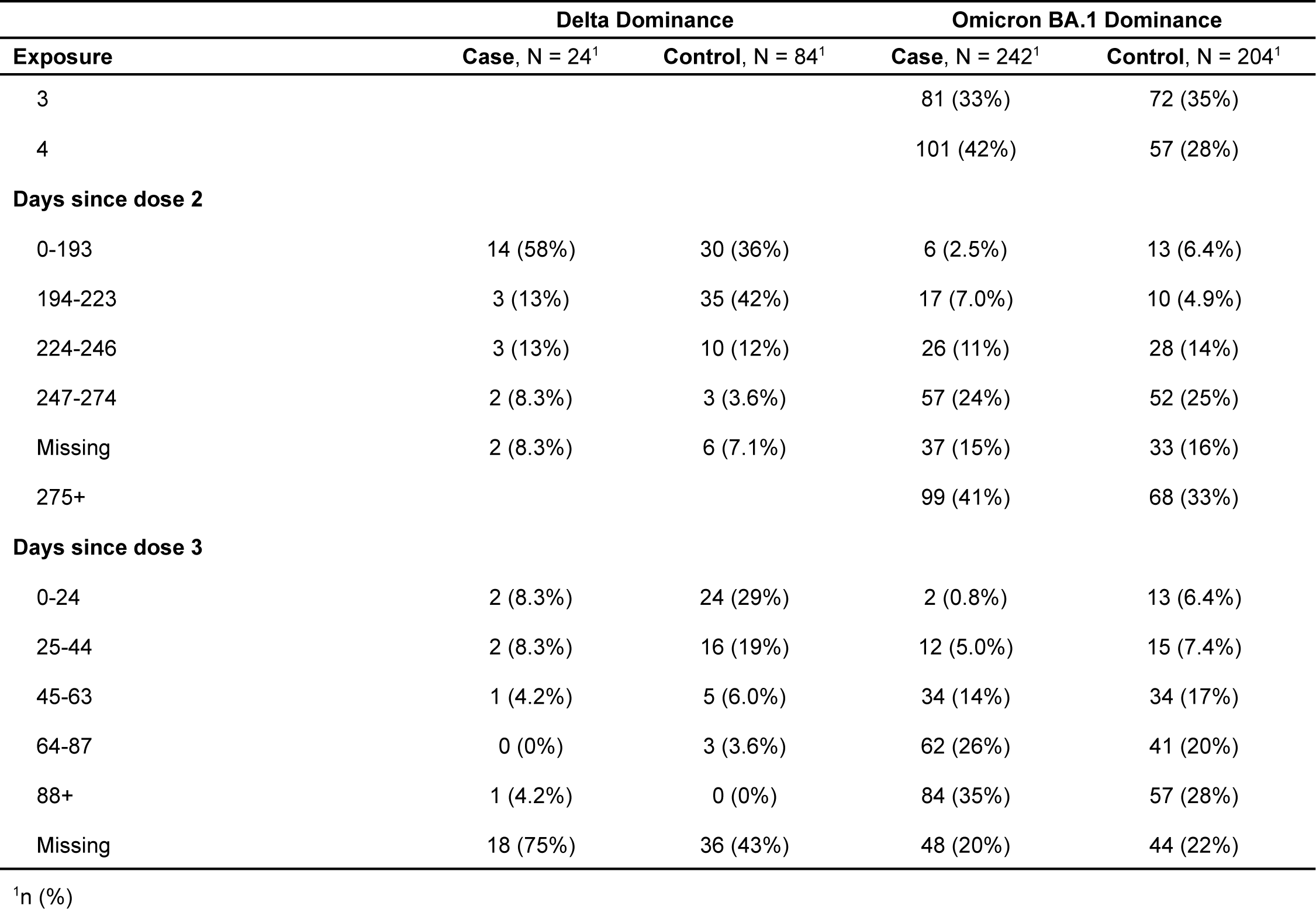
Baseline characteristics of the two subgroups, split by case/control status. Participants whose samples were successful on at least one of the three live virus assays were included in this summary.

During the Delta dominance period (N = 108), there were more women than men (58% vs 42%) and the largest age group was 45-64 years (44%), followed by 65+ years (29%). Most analysed participants were white British (90%) and most had ChAdOx1 (AstraZeneca) as their second vaccine dose (63%). 51% of participants had a booster vaccine dose before taking their PCR/LFD test. Most participants had their PCR/LFD test result recorded in the third fortnight of this dominance period (52%). Most participants took their PCR/LFD test within 193 days of having their second vaccine dose (41%). 24% of participants took their PCR/LFD test within 24 days of having their 3rd dose and 17% within 25-44 days.

During the Omicron BA.1 dominance period (N = 446), there were also more women than men (62% vs 38%) and the largest age group remained 45-64 years (49%), followed by 65+ years (34%). Likewise, most analysed participants were white British (91%) and had ChAdOx1 (AstraZeneca) as their second vaccine dose (51%). 79% of participants had a booster vaccine dose before taking their PCR/LFD test. Most participants had their PCR/LFD test result recorded in the fifth fortnight of this dominance period (35%). Most participants took their PCR/LFD test after 275+ days after having their second vaccine dose (37%). Most participants took their PCR/LFD test after 88+ days after having their booster dose (32%).

The differences in exposure stratum proportions between the two dominance periods reflect the likely temporal and variant-related changes in risk associated with these exposures.

Figure 2 shows the distributions of log_2_(IC50) values across the duration of the study, split by live virus assay variant and case/control status. These distributions indicate that, amongst the study participants, there was substantially greater neutralisation activity against the Delta variant compared to the Omicron BA.1 and BA.2 variants. Figures 3-5 show the distribution of log_2_(IC50) values split by assay variant, case/control status and variant dominance period.

**Figure 2.**
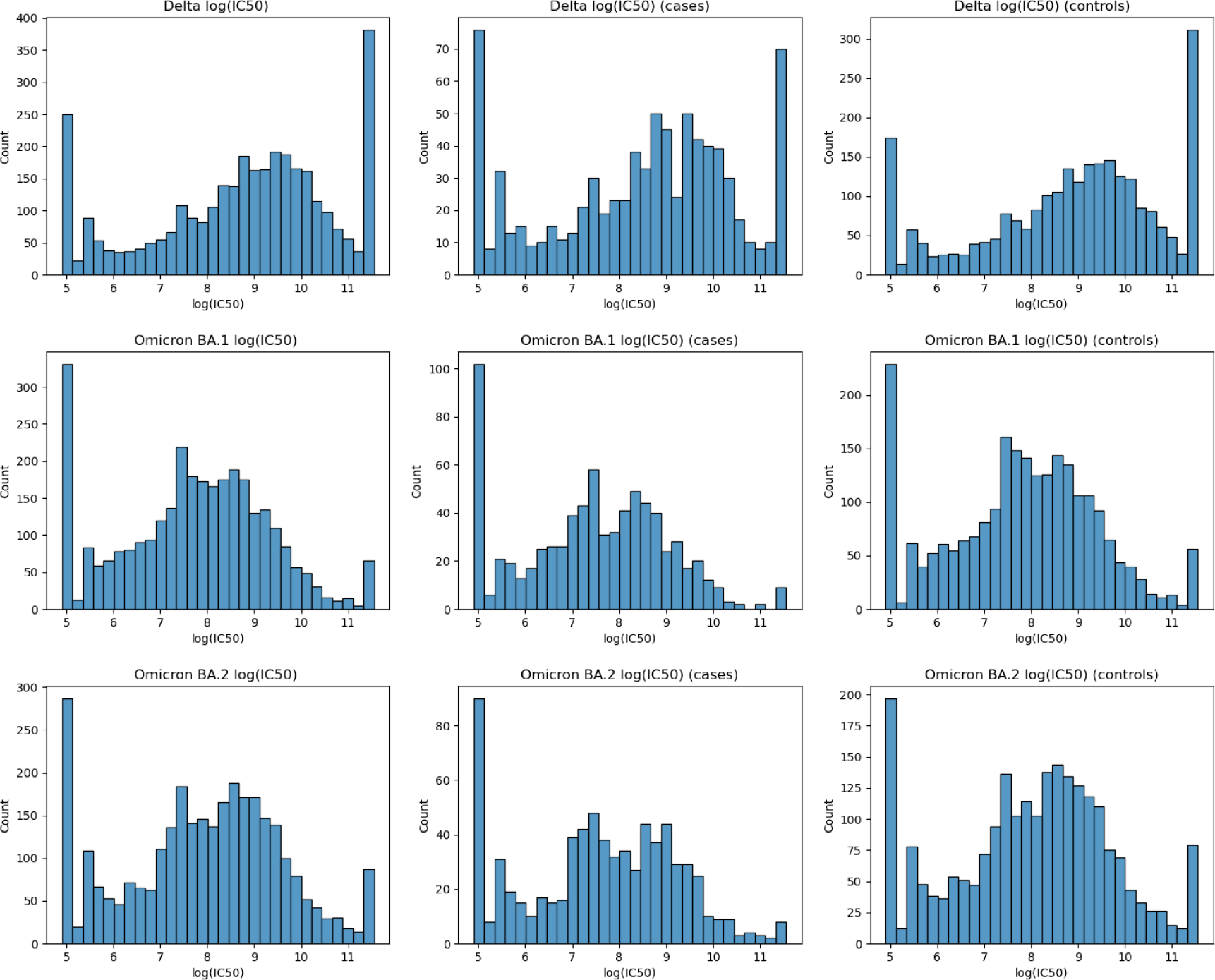
log_2_(IC50) values for the full duration of the study

**Figure 3.**
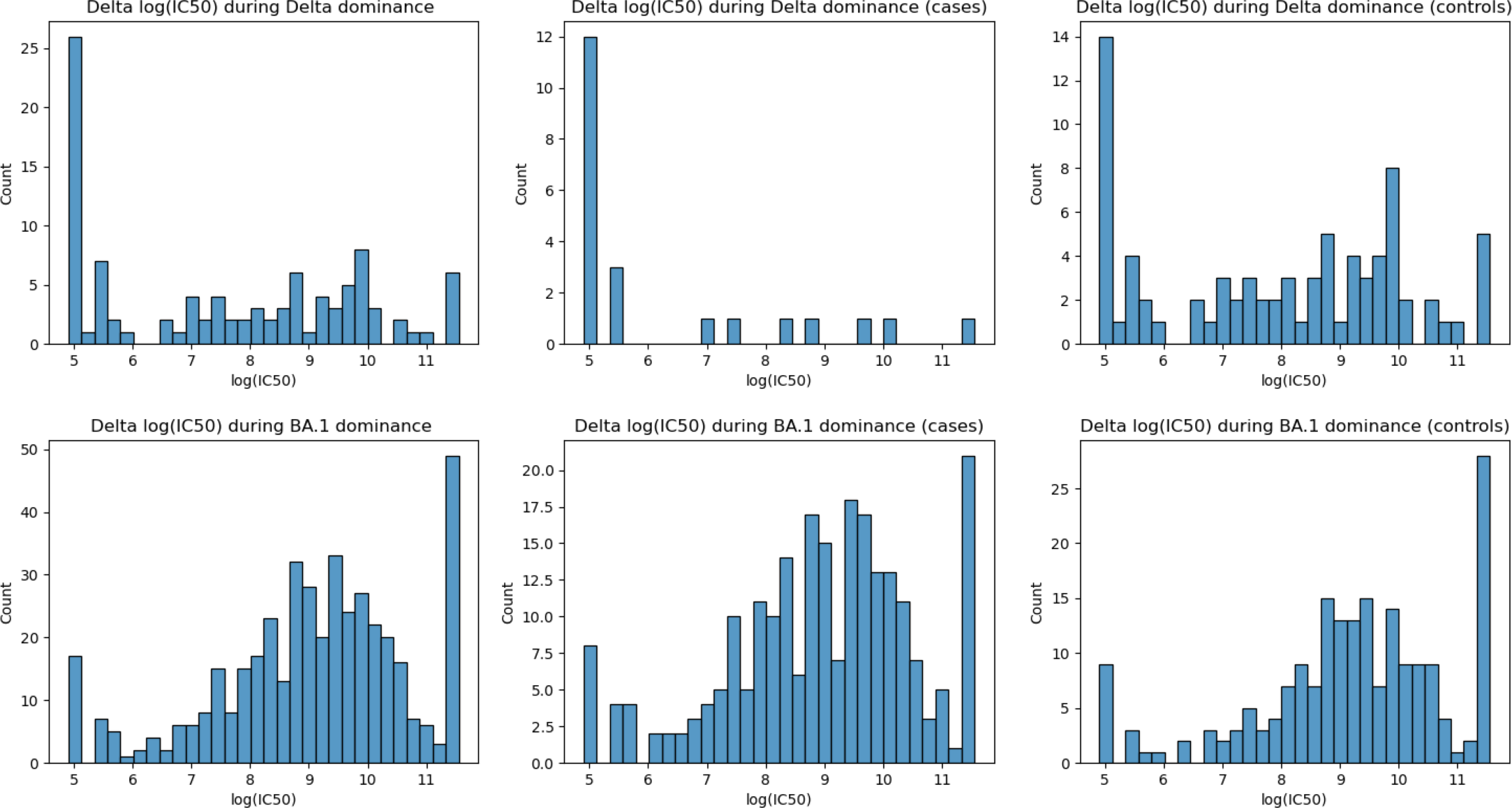
Delta log_2_(IC50) values

**Figure 4.**
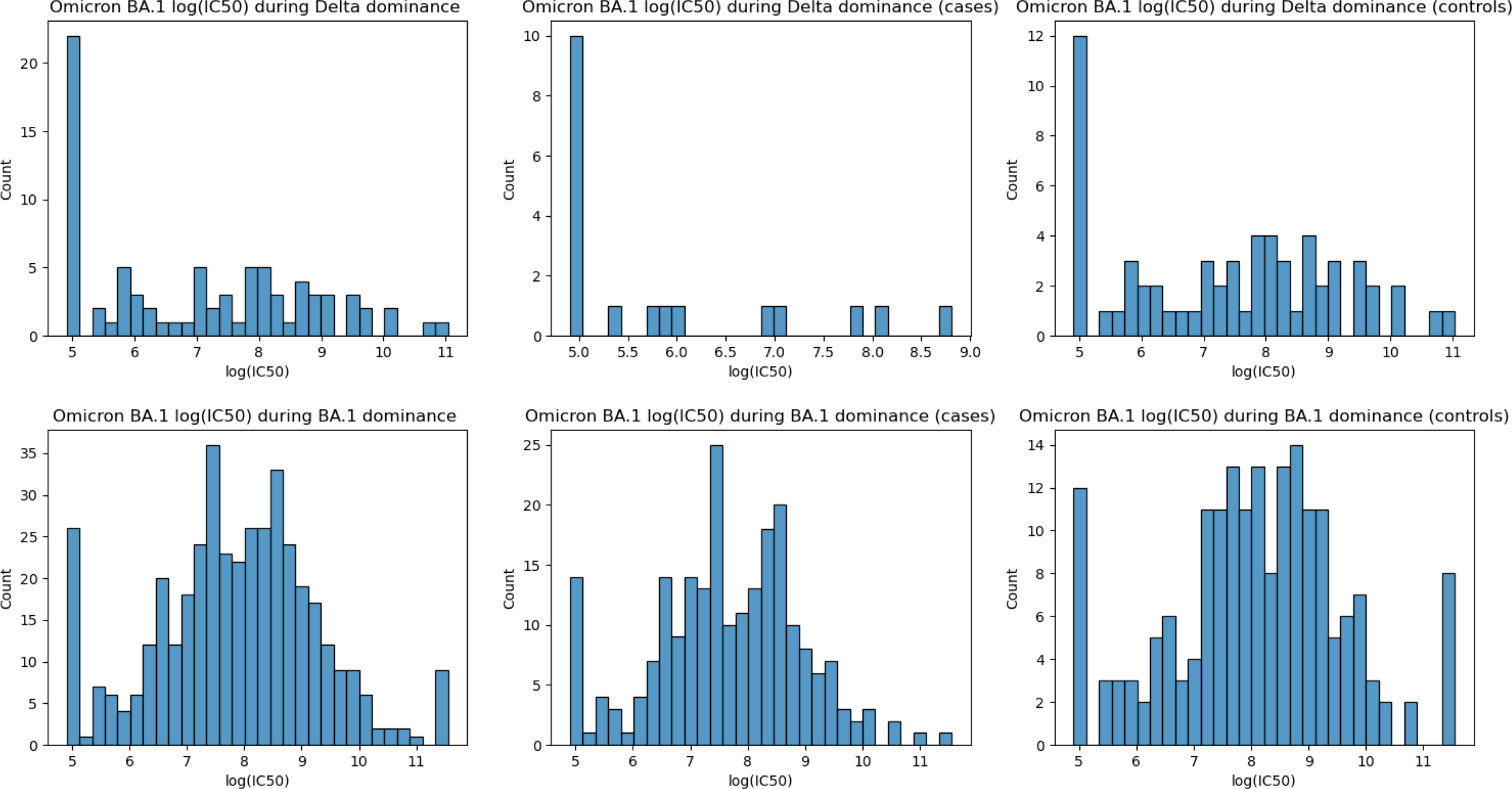
Omicron BA.1 log_2_(IC50) values

**Figure 5.**
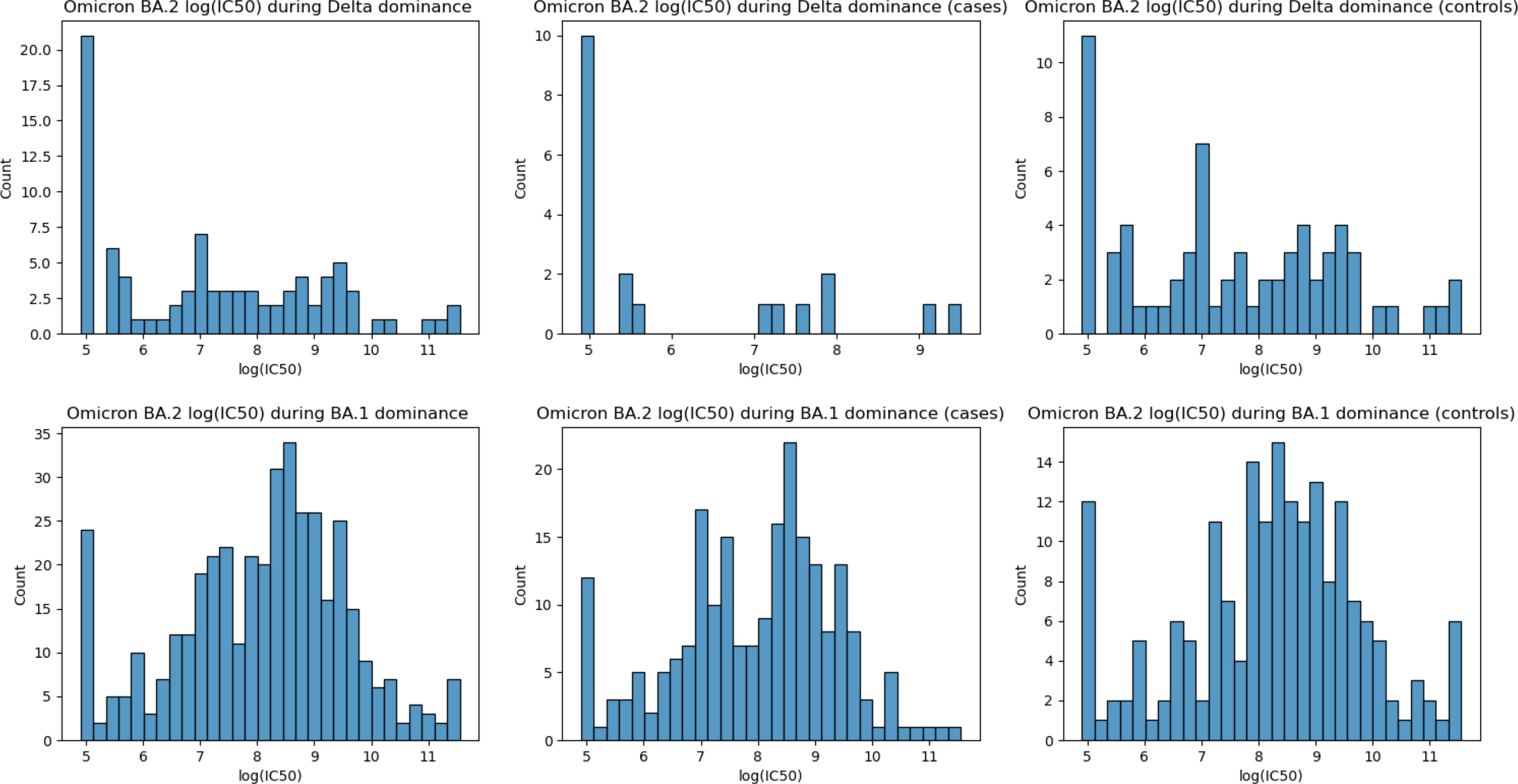
Omicron BA.2 log_2_(IC50) value

These figures demonstrate an increase in neutralisation activity against all three variants during the Omicron BA.1 dominance period compared with the Delta dominance period, in both cases and controls.

Appendix 1 shows mean log_2_(IC50) values for each variant, stratified by covariate categories and whether the individual had received a booster prior to taking their microsample test. Of note is the significantly greater average Delta log_2_(IC50) value in the BNT162b2 mRNA vaccine second dose recipients (8.65 (8.04-9.26)) compared with ChAdOx1 second dose vaccinees (7.05 (6.52-7.58)) during the Delta dominance period. However, this relationship does not hold for Omicron BA.1 and BA.2 average log_2_(IC50) values during the same period and is reversed during the Omicron BA.1 dominance period for Omicron BA.1 and BA.2 variants. Notable also are the significantly higher average log_2_(IC50) values for all variants in the 65+ year age group compared to the 25-44 and 45-65 year age groups during the Delta dominance period. During the Omicron BA.1 dominance period the 45-65 year age group had significantly higher average log_2_(IC50) values compared to the 65+ year age group for the Delta and Omicron BA.2 variants. During the latter period, there was weak evidence of participants who are not clinically vulnerable having increased average Delta variant log_2_(IC50) values compared to the clinically vulnerable. Having had a booster dose before taking the capillary microsample test significantly increased the average log_2_(IC50) values for all variants during the Delta dominance period. During the Omicron BA.1 period, there was no evidence of this increase for all variants.

Table 5 shows the crude infection odds ratios with 95% confidence intervals, p values and case/control numbers for each live virus assay. The adjusted odds ratios are reported in Table 6. During the Delta dominance period, a one unit increase in the log_2_(IC50) value (i.e. a doubling in the IC50 value) is associated with reduced risk of SARS-CoV-2 infection for all variants (Delta, crude OR = 0.67 (0.51-0.87), adjusted OR = 0.69 (0.50-0.95); Omicron BA.1, crude OR = 0.53 (0.36-0.79), adjusted OR = 0.56 (0.34-0.90); Omicron BA.2, crude OR = 0.61 (0.44-0.86), adjusted OR = 0.63 (0.43-0.93). During the Omicron BA.1 dominance period, only Omicron BA.1 neutralisation has a protective effect against infection (crude OR = 0.80 (0.70-0.92), adjusted OR = 0.80 (0.69-0.92)).

**Table 5.** Crude odds ratios.

| Variant | Time period (dominant variant) |  |
| --- | --- | --- |
|  | 01/11/21 - 06/12/21 (Delta) | 15/12/21-17/02/22 (BA.1) |
| **Delta log<sub>2</sub>(IC50)** | OR = 0.67 (0.51-0.87)<br>p = 0.003<br>Cases: 22<br>Controls: 80 | OR = 0.91 (0.81-1.02)<br>p = 0.111<br>Cases: 238<br>Controls: 198 |
| **BA.1 log<sub>2</sub>(IC50)** | OR = 0.53 (0.36-0.79)<br>p = 0.002<br>Cases: 19<br>Controls: 63 | OR = 0.80 (0.70-0.92)<br>p = 0.002<br>Cases: 224<br>Controls: 190 |
| **BA.2 log<sub>2</sub>(IC50)** | OR = 0.61 (0.44-0.86)<br>p = 0.005<br>Cases: 20<br>Controls: 69 | OR = 0.88 (0.77-1.01)<br>p = 0.064<br>Cases: 218<br>Controls: 189 |

**Table 6.** Adjusted odds ratios (adjusted for age, sex, clinical vulnerability, and booster dose status at the time of PCR/LFD test)

| Variant | Time period (dominant variant) |  |
| --- | --- | --- |
|  | 01/11/21 - 06/12/21 (Delta) | 15/12/21-17/02/22 (BA.1) |
| **Delta log<sub>2</sub>(IC50)** | OR = 0.69 (0.50-0.95)<br>p = 0.024<br>Cases: 22<br>Controls: 80 | OR = 0.91 (0.80-1.02)<br>p = 0.099<br>Cases: 238<br>Controls: 198 |
| **BA.1 log<sub>2</sub>(IC50)** | OR = 0.56 (0.34-0.90)<br>p = 0.018<br>Cases: 19<br>Controls: 63 | OR = 0.80 (0.69-0.92)<br>p = 0.002<br>Cases: 224<br>Controls: 190 |
| **BA.2 log<sub>2</sub>(IC50)** | OR = 0.63 (0.43-0.93)<br>p = 0.019<br>Cases: 20<br>Controls: 69 | OR = 0.88 (0.77-1.01)<br>p = 0.065<br>Cases: 218<br>Controls: 189 |

Appendix 2 shows protection curves for each live virus variant and dominance period. There is a general trend towards greater protection against infection for higher log_2_(IC50) quintiles, with the top quintile of Omicron BA.1 log_2_(IC50) offering the greatest protection during the Delta dominance period (OR = 0.07 (0.01-0.88)) and Omicron BA.1 dominance period (OR 0.44 (0.23-0.83)).

Appendix 3 shows the effect of the gap duration between dominance periods on the adjusted odds ratios. During the Delta dominance period, longer gaps result in lower odds ratios for all three variants.The odds ratios remain largely unchanged during the Omicron BA.1 period.

Appendix 4 shows how log_2_(IC50) values changed over time after receiving a booster dose and before testing positive for SARS-CoV-2. During the Delta dominance period the number of samples was too small for meaningful analysis. During the Omicron BA.1. period, post-booster log_2_(IC50) values exhibit waning over time for all three variants. For all variants, log_2_(IC50) values tend to be lower closer to the date of a positive test.

## Discussion

We found that greater live virus neutralisation activity in an individual’s microsample was associated with a reduced risk of that individual developing a subsequent SARS-CoV-2 infection, and our results highlight the potential robustness of this methodological approach. We found greater inhibition of Omicron BA.1 live virus was associated with a reduction in infection risk during both the Delta and Omicron BA.1 dominance periods. Delta virus inhibition was associated with infection risk reduction during the Delta dominance period, but we found no association between Delta inhibition and protection against infection during the Omicron BA.1 dominance period. These results are consistent with earlier findings suggesting that immunity against Omicron BA.1 is protective against Delta, but not vice versa (20,21).

We found significantly greater neutralisation activity against Delta in BNT162b2 mRNA second dose vaccinees compared with ChAdOx1 second dose recipients during the Delta dominance period. This is consistent with earlier findings that show that a two-dose BNT162b2 vaccine course elicits an anti-spike antibody response that is an order of magnitude greater than that of ChAdOx1 (7). We found no evidence of this difference in neutralisation for Omicron BA.1 and BA.2 in the same period. This finding corroborates earlier comparisons between two-dose regimens of the two vaccines, which demonstrate comparable efficacy of both against Omicron (22). We also observed that during the Omicron BA.1 dominance period, ChAdOx1 second dose recipients had significantly greater inhibition of all three variants. This may be explained by the combination of younger participants being offered booster doses closer to the Omicron BA.1 dominance period (23), and the greater frequency of ChAdOx1 vaccination in this age group. The prioritisation of booster doses for older participants during the Delta dominance period may also explain the significantly greater inhibition of all variants in the 65+ year old age group during that period (24).

The ability to investigate variant-specific NAb titres as correlates of protection against SARS-CoV-2 infection before and after substantial antigenic shift of the viral genome are both a major strength and novelty of this study. Previous studies have investigated the relationships between variant-specific NAbTs and vaccinations in different population subgroups (19,25–27) and with reinfections as compared to primary infections (8).

An important limitation of this study is the issue of time-varying confounding. The change in the dominant variant and the subsequent population-wide rollout of booster vaccines occurring during the study make it challenging to draw unbiased comparisons between participants across time. The start and end dates of variant dominance could only be approximately estimated, making it difficult to reliably attribute a given infection event to the causative variant. Future research should incorporate genomic sequencing of infections to address this problem. As a consequence of having to split the study participants into subgroups to address time-varying confounding, this study was insufficiently powered to adjust infection risk estimates by time since last vaccine dose, peak nAb titres post vaccine, and second dose and booster vaccine types. These are important immunological covariates that should be included in the analyses in future studies.

Boosters have the potential to drastically alter the immunity landscape in the population. However, we did not find evidence of boosters substantially changing neutralisation levels during the Omicron BA.1 dominance period, suggesting that many participants may have experienced asymptomatic infections during that time. Future work should address the interaction between variant-specific virus inhibition and vaccination status as related to the risk of breakthrough infections.

We have shown that collecting blood from individuals using capillary microsamples can be performed at scale and we have demonstrated the feasibility of using live virus neutralisation assays with such microsamples. Greater inhibition of Omicron BA.1 was associated with a reduction in infection risk during both the Delta and Omicron BA.1 dominance periods. Delta virus inhibition was only associated with infection risk reduction during the Delta dominance period and not during the Omicron BA.1 period. Our findings suggest that variant-specific serosurveillance of immunity and protection against SARS-CoV-2 infection at the population level could inform public health policy in near-real time.

## Data availability

Given the sensitive content in our dataset (information on health) for this study, we cannot publish data at the individual level, publicly. We are sharing individual record level data (excluding any data or variables originating from linkage via NHS Digital) on the ONS SRS (DOI: 10.57906/s5f5-nq13). The data are available under restricted access and can be obtained by submitting a request directly to the SRS. We regularly share results and updates on the study via a “Findings so far” section on our website - https://ucl-virus-watch.net/ This study uses NHS HES (Admitted Patient Care, Outpatient Bookings, Emergency Care Dataset and Critical Care), Vaccination (NIMS), and COVID-19 testing data (Pillar 2 and SGSS) that were provided within the terms of a data-sharing agreement (DARS-NIC-372269-N8D7Z-V1.6) to the researchers by the Health and Social Care Information Centre (NHS Digital). The data do not belong to the authors and may not be shared by the authors, except in aggregate form for publication. Data can be obtained by submitting a data request through the NHS Digital Data Access Request Service.

## Author contributions

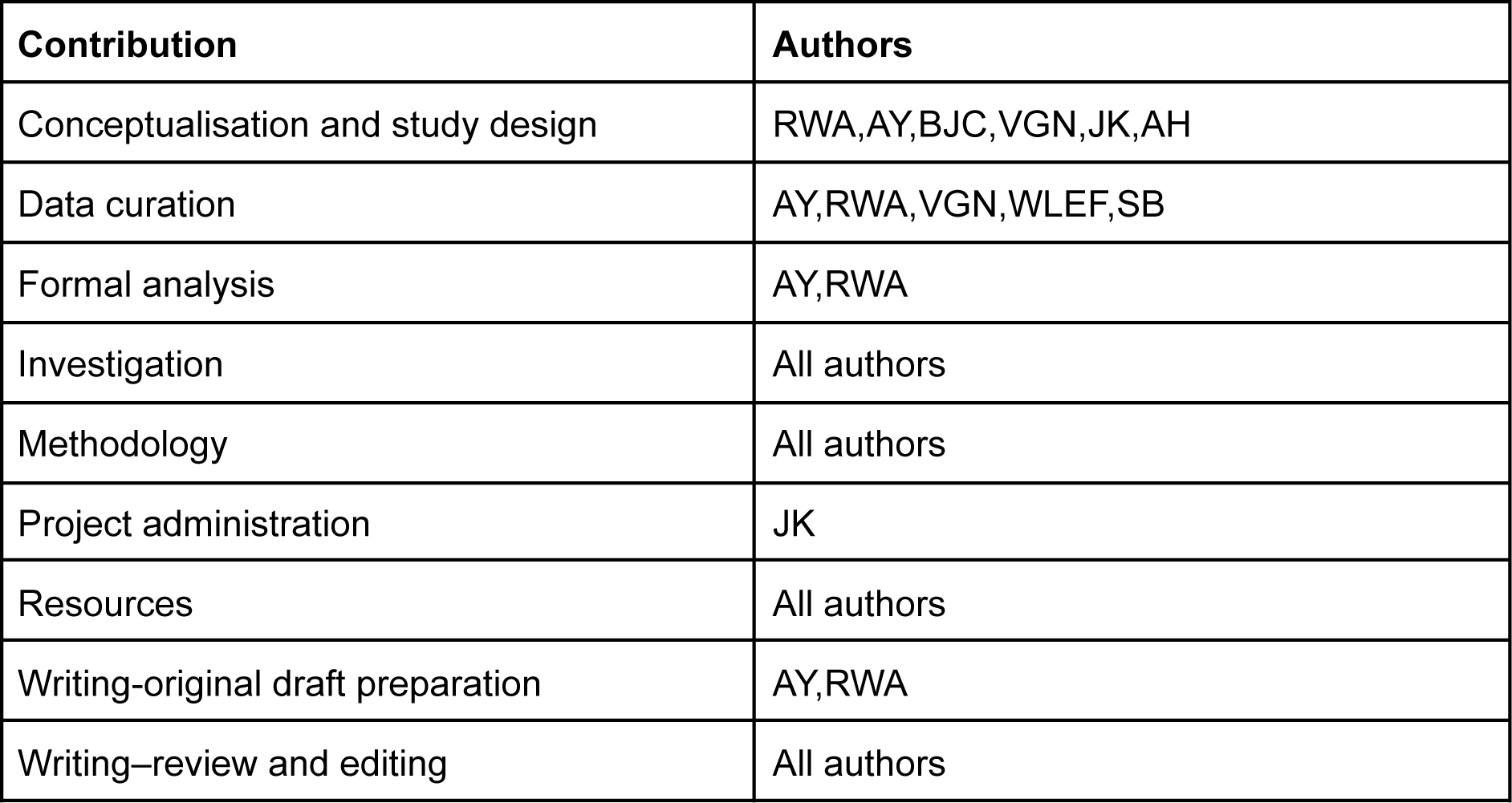

Distribution of log_2_(IC50) values by case/control status, live virus assay variant and dominant variant time period

## Appendix 1. Mean log2(IC50) values by variant, baseline characteristics and time period

ANOVA one way test p-value is reported next to the covariate heading and if significant differences were found, the result of Tukey post-hoc tests with p<0.05 are reported in brackets

Period 1: 01/11/21 - 6/12/21 (Delta dominance)

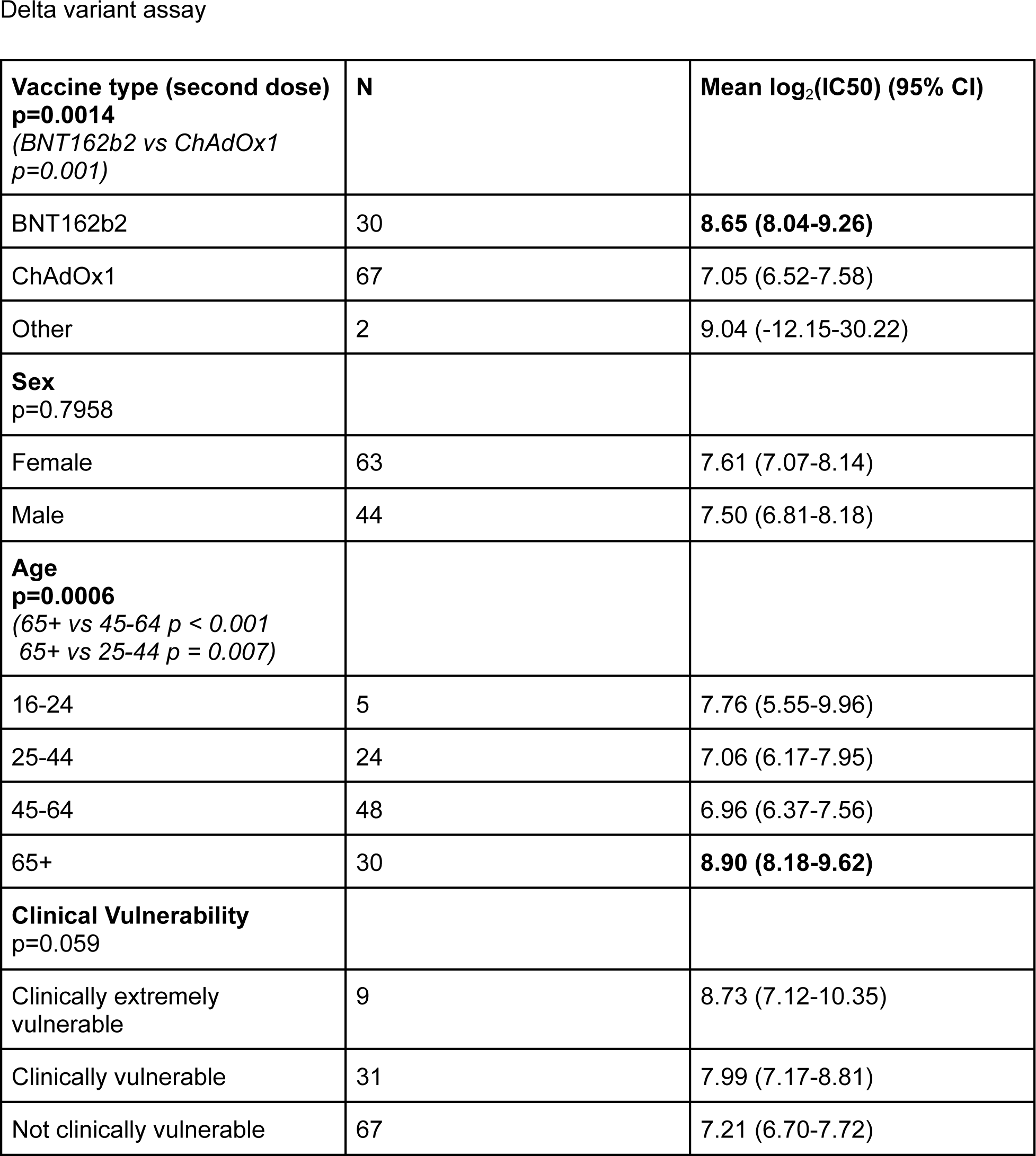
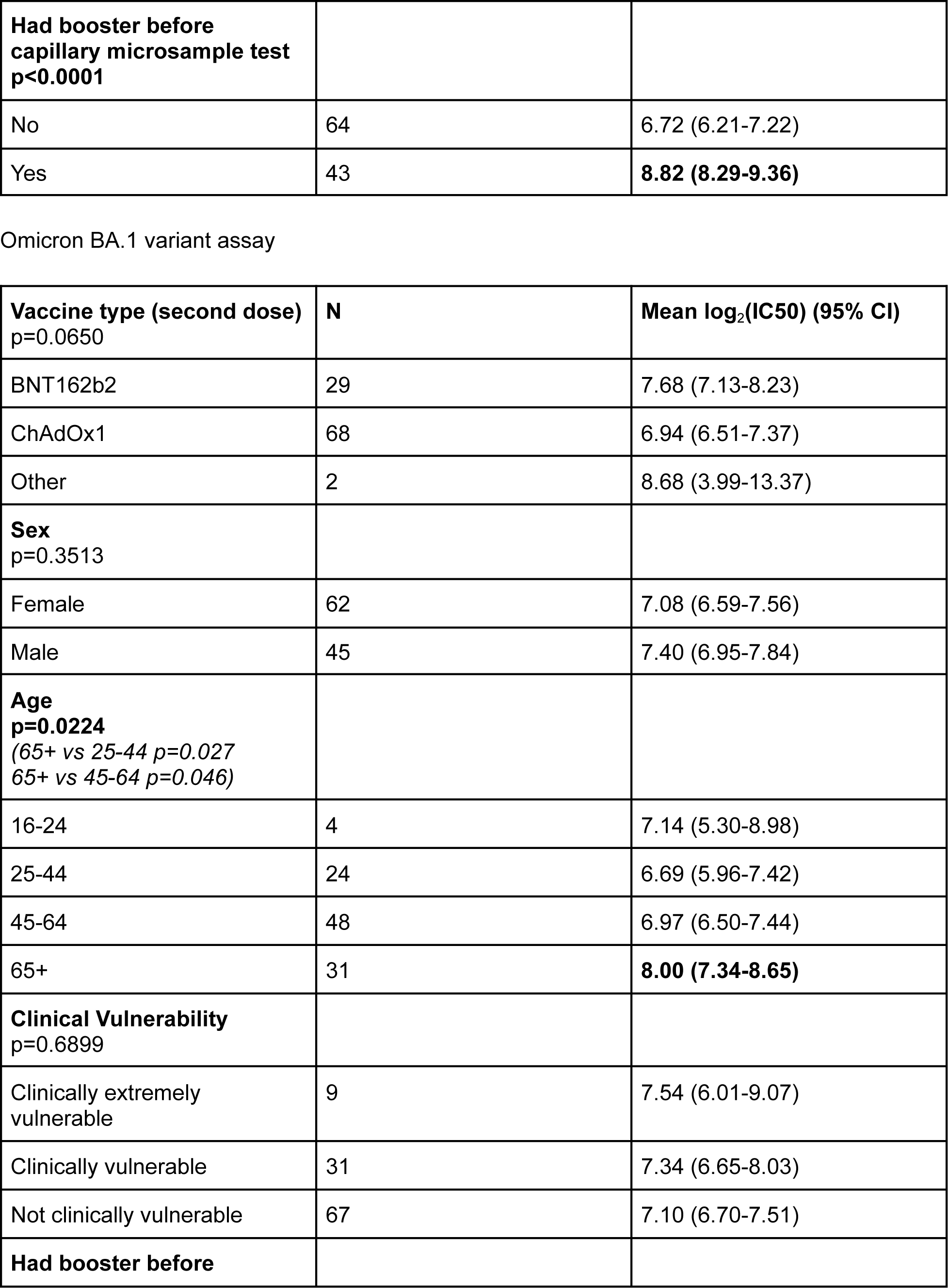
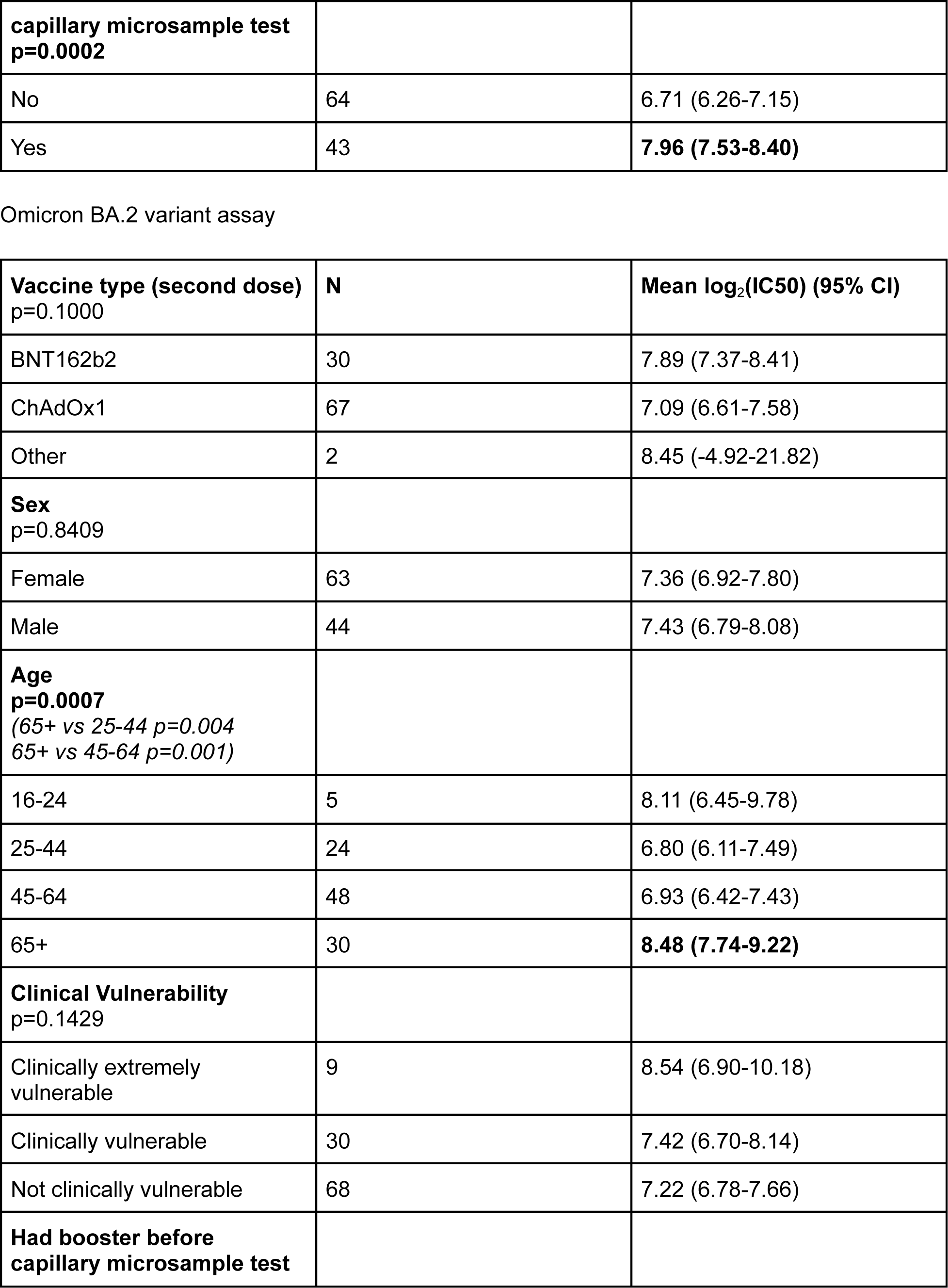
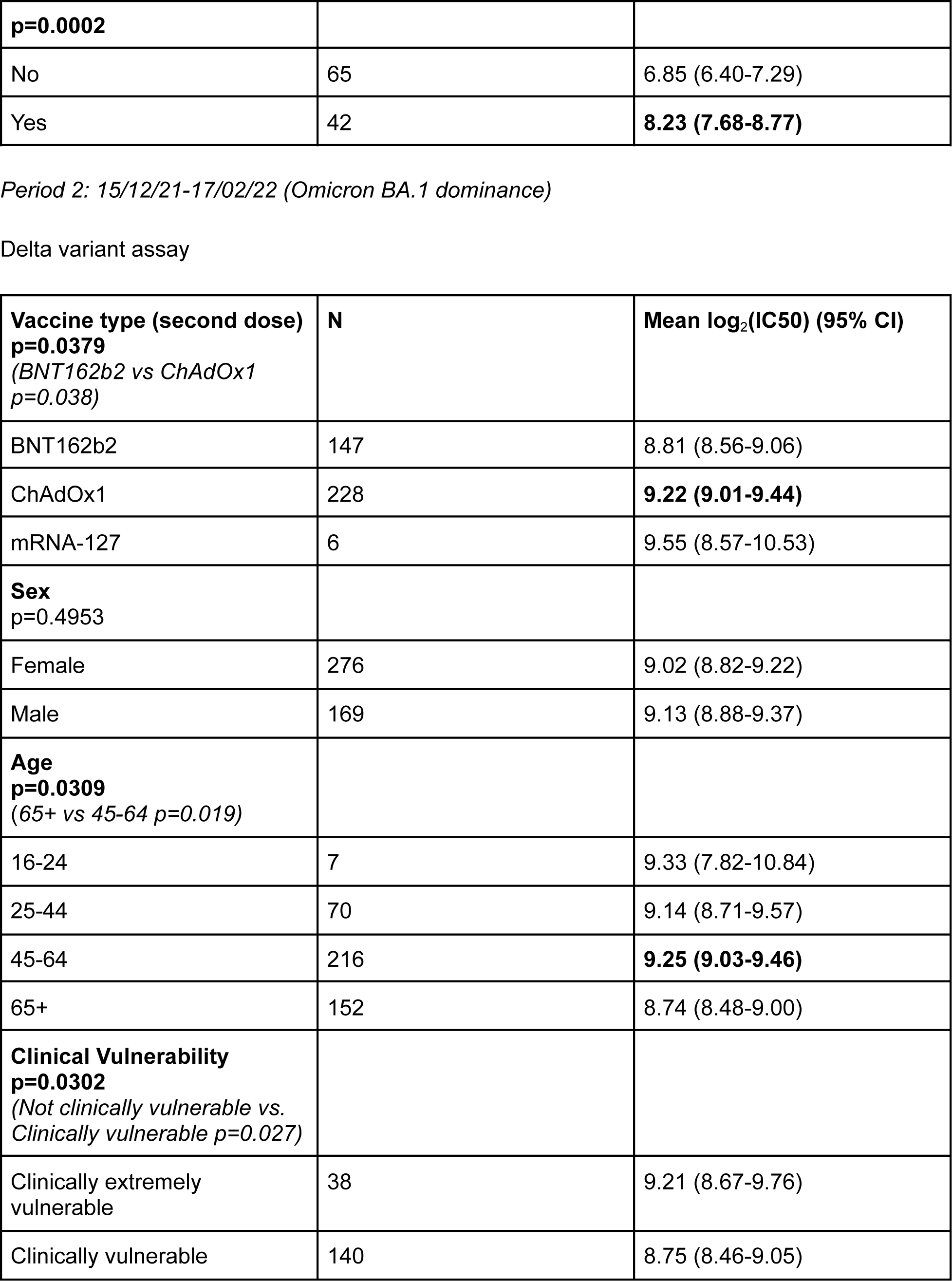
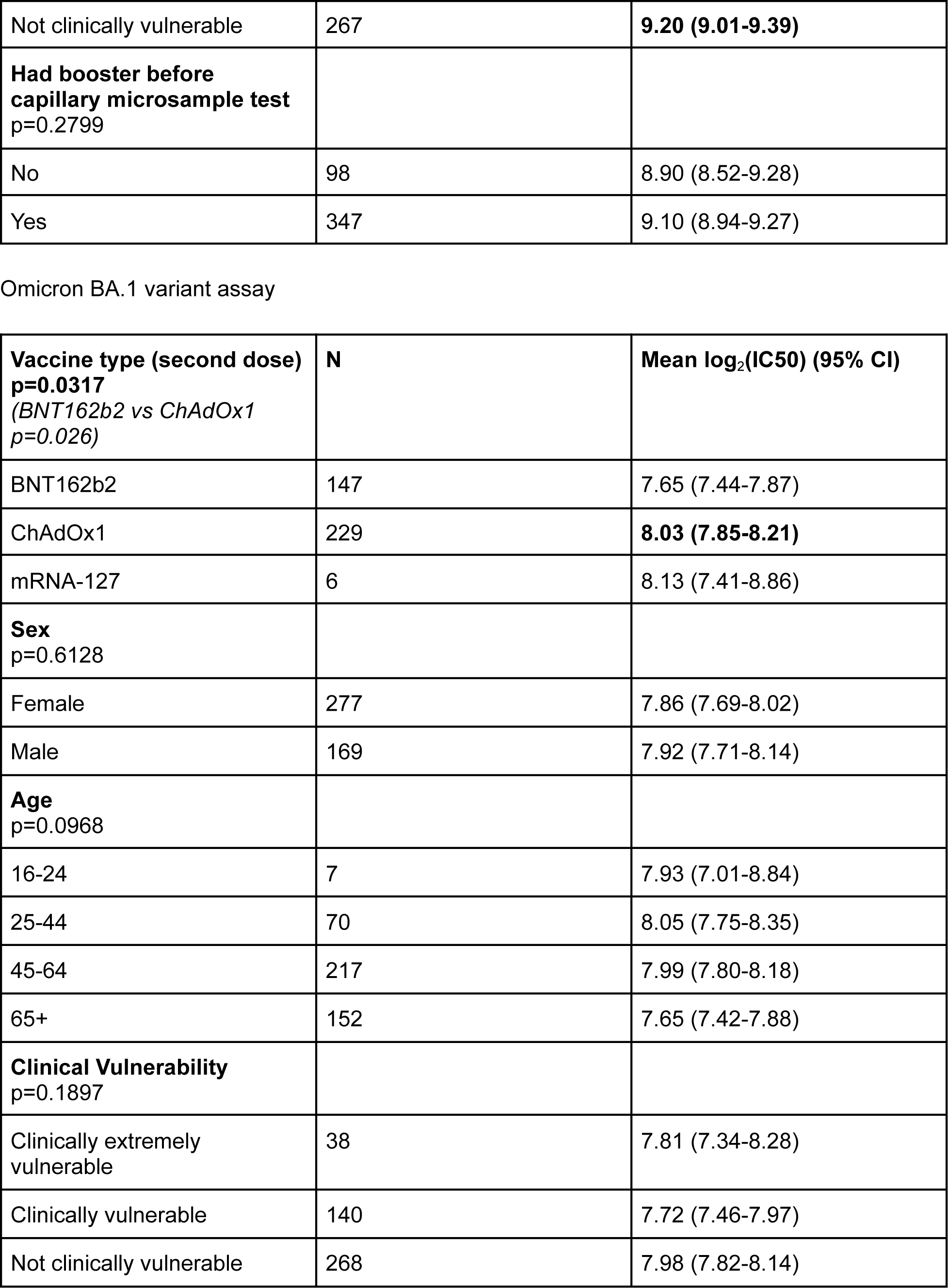
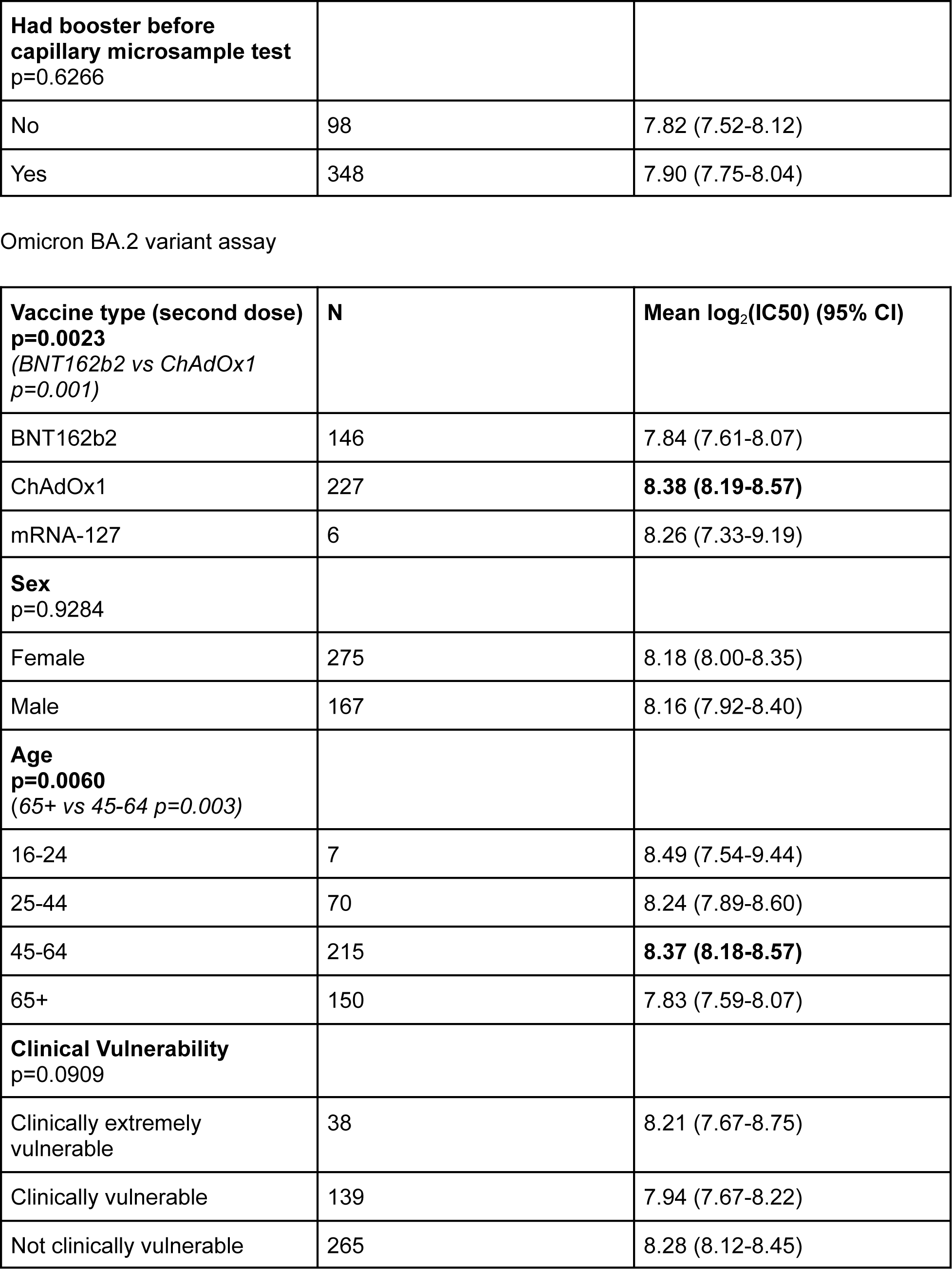
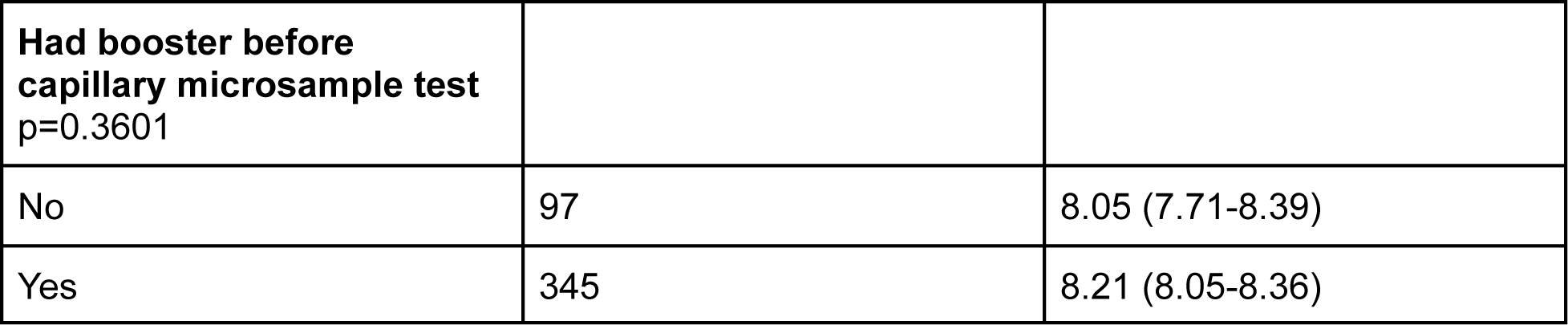

| **Vaccine type (second dose)**<br>**p=0.0014**<br>(BNT162b2 vs ChAdOx1<br>$p=0.001$) | **N** | **Mean $\log_2$(IC50) (95% CI)** |
| --- | --- | --- |
| BNT162b2 | 30 | **8.65 (8.04-9.26)** |
| ChAdOx1 | 67 | 7.05 (6.52-7.58) |
| Other | 2 | 9.04 (-12.15-30.22) |
| **Sex**<br>**p=0.7958** |  |  |
| Female | 63 | 7.61 (7.07-8.14) |
| Male | 44 | 7.50 (6.81-8.18) |
| **Age**<br>**p=0.0006**<br>(65+ vs 45-64 $p < 0.001$<br>65+ vs 25-44 $p = 0.007$) | | |
| 16-24 | 5 | 7.76 (5.55-9.96) |
| 25-44 | 24 | 7.06 (6.17-7.95) |
| 45-64 | 48 | 6.96 (6.37-7.56) |
| 65+ | 30 | **8.90 (8.18-9.62)** |
| **Clinical Vulnerability**<br>**p=0.059** |  |  |
| Clinically extremely vulnerable | 9 | 8.73 (7.12-10.35) |
| Clinically vulnerable | 31 | 7.99 (7.17-8.81) |
| Not clinically vulnerable | 67 | 7.21 (6.70-7.72) |
| **Had booster before capillary microsample test**<br>**p<0.0001** |  |  |
| No | 64 | 6.72 (6.21-7.22) |
| Yes | 43 | **8.82 (8.29-9.36)** |
| Omicron BA.1 variant assay |  |  |
| **Vaccine type (second dose)**<br>p=0.0650 | **N** | **Mean log<sub>2</sub>(IC50) (95% CI)** |
| BNT162b2 | 29 | 7.68 (7.13-8.23) |
| ChAdOx1 | 68 | 6.94 (6.51-7.37) |
| Other | 2 | 8.68 (3.99-13.37) |
| **Sex**<br>p=0.3513 |  |  |
| Female | 62 | 7.08 (6.59-7.56) |
| Male | 45 | 7.40 (6.95-7.84) |
| **Age**<br>**p=0.0224**<br>(65+ vs 25-44 p=0.027<br>65+ vs 45-64 p=0.046) |  |  |
| 16-24 | 4 | 7.14 (5.30-8.98) |
| 25-44 | 24 | 6.69 (5.96-7.42) |
| 45-64 | 48 | 6.97 (6.50-7.44) |
| 65+ | 31 | **8.00 (7.34-8.65)** |
| **Clinical Vulnerability**<br>p=0.6899 |  |  |
| Clinically extremely vulnerable | 9 | 7.54 (6.01-9.07) |
| Clinically vulnerable | 31 | 7.34 (6.65-8.03) |
| Not clinically vulnerable | 67 | 7.10 (6.70-7.51) |
| **Had booster before** |  |  |
| **capillary microsample test**<br>**p=0.0002** |  |  |
| No | 64 | 6.71 (6.26-7.15) |
| Yes | 43 | **7.96 (7.53-8.40)** |
| Omicron BA.2 variant assay |  |  |
| **Vaccine type (second dose)**<br>**p=0.1000** | **N** | **Mean log<sub>2</sub>(IC50) (95% CI)** |
| BNT162b2 | 30 | 7.89 (7.37-8.41) |
| ChAdOx1 | 67 | 7.09 (6.61-7.58) |
| Other | 2 | 8.45 (-4.92-21.82) |
| **Sex**<br>**p=0.8409** |  |  |
| Female | 63 | 7.36 (6.92-7.80) |
| Male | 44 | 7.43 (6.79-8.08) |
| **Age**<br>**p=0.0007**<br>(65+ vs 25-44 p=0.004<br>65+ vs 45-64 p=0.001) |  |  |
| 16-24 | 5 | 8.11 (6.45-9.78) |
| 25-44 | 24 | 6.80 (6.11-7.49) |
| 45-64 | 48 | 6.93 (6.42-7.43) |
| 65+ | 30 | **8.48 (7.74-9.22)** |
| **Clinical Vulnerability**<br>**p=0.1429** |  |  |
| Clinically extremely vulnerable | 9 | 8.54 (6.90-10.18) |
| Clinically vulnerable | 30 | 7.42 (6.70-8.14) |
| Not clinically vulnerable | 68 | 7.22 (6.78-7.66) |
| **Had booster before capillary microsample test** |  |  |
| **p=0.0002** |  |  |
| No | 65 | 6.85 (6.40-7.29) |
| Yes | 42 | **8.23 (7.68-8.77)** |
| **Vaccine type (second dose)**<br>**p=0.0379**<br>(BNT162b2 vs ChAdOx1<br>*p=0.038*) | **N** | **Mean log<sub>2</sub>(IC50) (95% CI)** |
| BNT162b2 | 147 | 8.81 (8.56-9.06) |
| ChAdOx1 | 228 | **9.22 (9.01-9.44)** |
| mRNA-127 | 6 | 9.55 (8.57-10.53) |
| **Sex**<br>**p=0.4953** |  |  |
| Female | 276 | 9.02 (8.82-9.22) |
| Male | 169 | 9.13 (8.88-9.37) |
| **Age**<br>**p=0.0309**<br>(65+ vs 45-64 *p=0.019*) |  |  |
| 16-24 | 7 | 9.33 (7.82-10.84) |
| 25-44 | 70 | 9.14 (8.71-9.57) |
| 45-64 | 216 | **9.25 (9.03-9.46)** |
| 65+ | 152 | 8.74 (8.48-9.00) |
| **Clinical Vulnerability**<br>**p=0.0302**<br>(Not clinically vulnerable vs.<br>Clinically vulnerable *p=0.027*) |  |  |
| Clinically extremely vulnerable | 38 | 9.21 (8.67-9.76) |
| Clinically vulnerable | 140 | 8.75 (8.46-9.05) |
| Not clinically vulnerable | 267 | **9.20 (9.01-9.39)** |
| **Had booster before capillary microsample test**<br>p=0.2799 |  |  |
| No | 98 | 8.90 (8.52-9.28) |
| Yes | 347 | 9.10 (8.94-9.27) |
| Omicron BA.1 variant assay |  |  |
| **Vaccine type (second dose)**<br>**p=0.0317**<br>(BNT162b2 vs ChAdOx1<br>p=0.026) | **N** | **Mean log<sub>2</sub>(IC50) (95% CI)** |
| BNT162b2 | 147 | 7.65 (7.44-7.87) |
| ChAdOx1 | 229 | **8.03 (7.85-8.21)** |
| mRNA-127 | 6 | 8.13 (7.41-8.86) |
| **Sex**<br>p=0.6128 |  |  |
| Female | 277 | 7.86 (7.69-8.02) |
| Male | 169 | 7.92 (7.71-8.14) |
| **Age**<br>p=0.0968 |  |  |
| 16-24 | 7 | 7.93 (7.01-8.84) |
| 25-44 | 70 | 8.05 (7.75-8.35) |
| 45-64 | 217 | 7.99 (7.80-8.18) |
| 65+ | 152 | 7.65 (7.42-7.88) |
| **Clinical Vulnerability**<br>p=0.1897 |  |  |
| Clinically extremely vulnerable | 38 | 7.81 (7.34-8.28) |
| Clinically vulnerable | 140 | 7.72 (7.46-7.97) |
| Not clinically vulnerable | 268 | 7.98 (7.82-8.14) |
| **Had booster before capillary microsample test**<br>p=0.6266 |  |  |
| No | 98 | 7.82 (7.52-8.12) |
| Yes | 348 | 7.90 (7.75-8.04) |
| Omicron BA.2 variant assay |  |  |
| **Vaccine type (second dose)**<br>**p=0.0023**<br>(BNT162b2 vs ChAdOx1<br>p=0.001) | **N** | **Mean log<sub>2</sub>(IC50) (95% CI)** |
| BNT162b2 | 146 | 7.84 (7.61-8.07) |
| ChAdOx1 | 227 | **8.38 (8.19-8.57)** |
| mRNA-127 | 6 | 8.26 (7.33-9.19) |
| **Sex**<br>p=0.9284 |  |  |
| Female | 275 | 8.18 (8.00-8.35) |
| Male | 167 | 8.16 (7.92-8.40) |
| **Age**<br>**p=0.0060**<br>(65+ vs 45-64 p=0.003) |  |  |
| 16-24 | 7 | 8.49 (7.54-9.44) |
| 25-44 | 70 | 8.24 (7.89-8.60) |
| 45-64 | 215 | **8.37 (8.18-8.57)** |
| 65+ | 150 | 7.83 (7.59-8.07) |
| **Clinical Vulnerability**<br>p=0.0909 |  |  |
| Clinically extremely vulnerable | 38 | 8.21 (7.67-8.75) |
| Clinically vulnerable | 139 | 7.94 (7.67-8.22) |
| Not clinically vulnerable | 265 | 8.28 (8.12-8.45) |
| **Had booster before**<br>**capillary microsample test**<br>p=0.3601 |  |  |
| No | 97 | 8.05 (7.71-8.39) |
| Yes | 345 | 8.21 (8.05-8.36) |

## Appendix 2. Log2(IC50) protection curves by variant and time period

**Figure 6.**
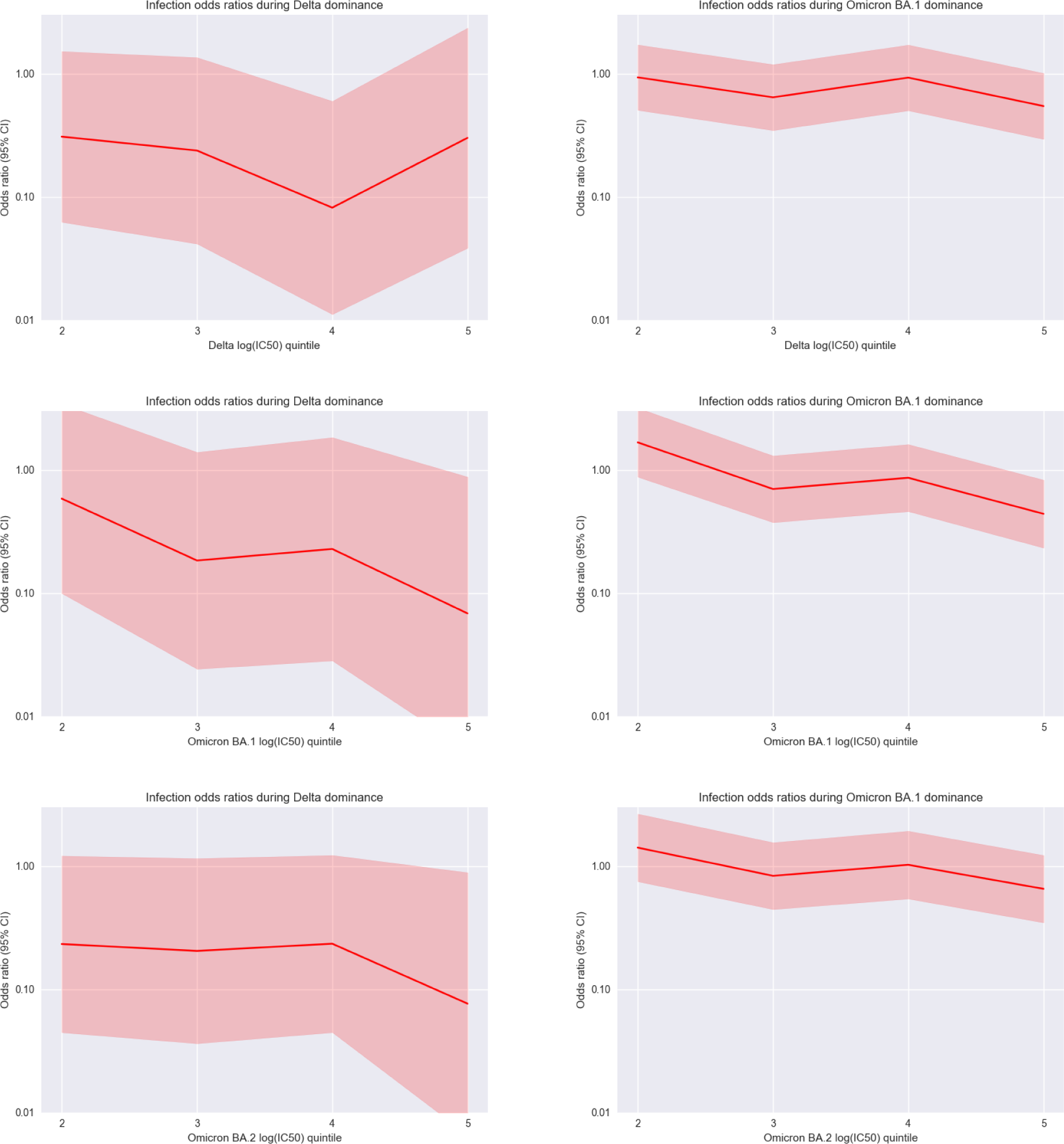
Protection curves

## Appendix 3. Sensitivity analyses

**Figure 7.**
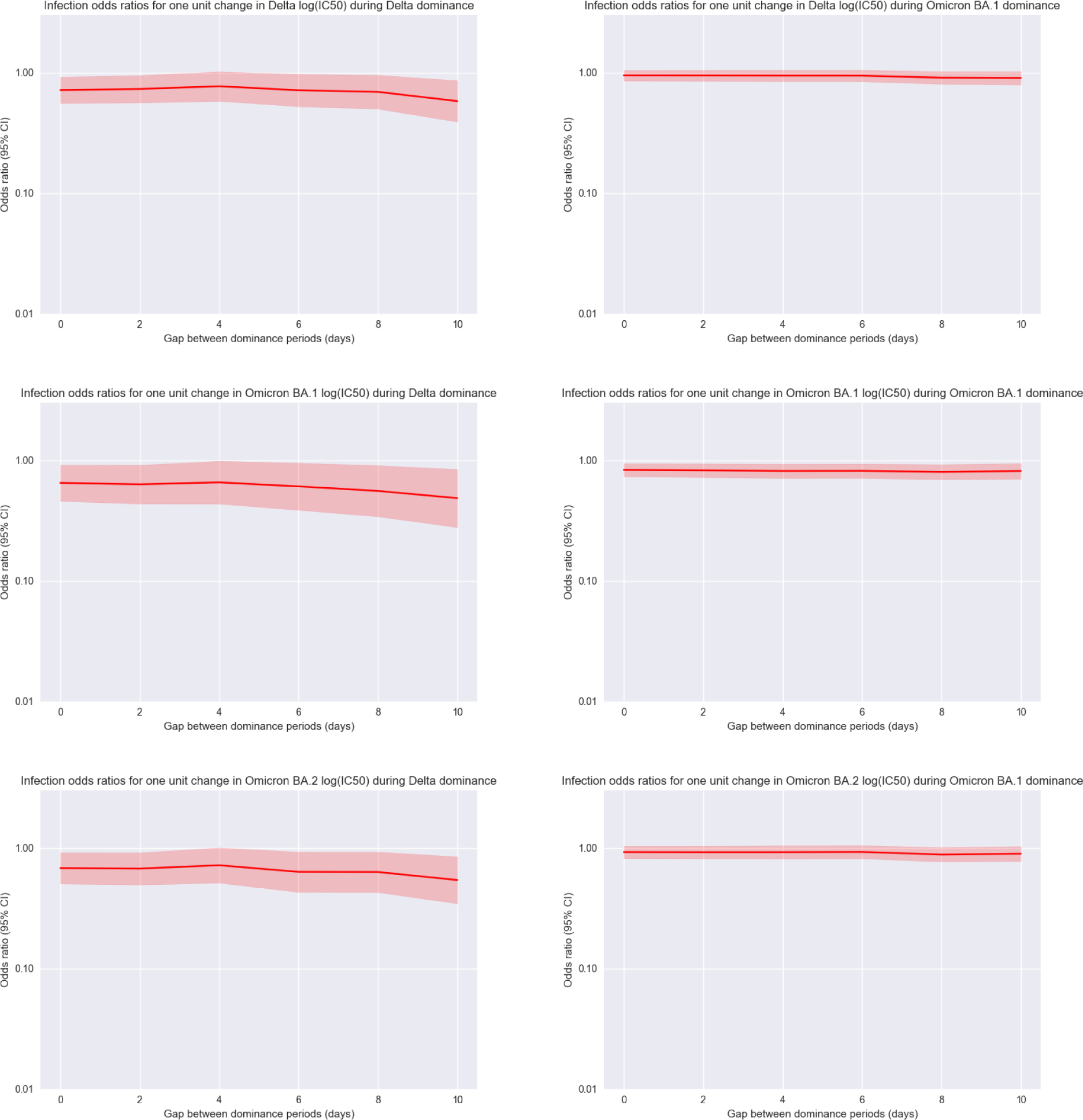
The impact of choosing different gap durations between dominance periods on infection odds ratios. The x axis represents the total number of days between the Delta and Omicron BA.1 dominance periods (eight days was the value used for generating all results in this study).

**Figure 8.**
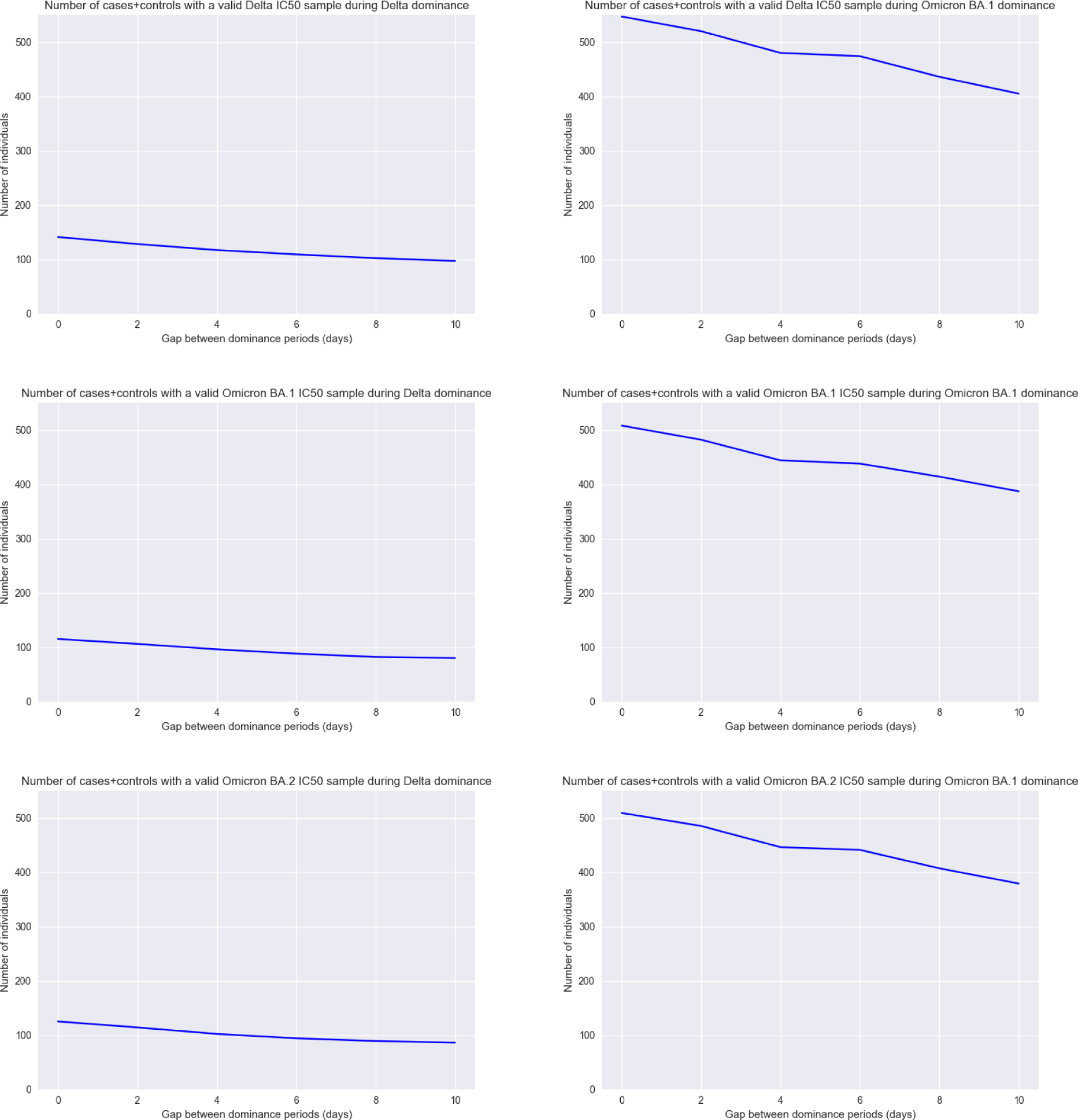
The impact of choosing different gap durations between dominance periods on the number of cases and controls for each live neutralisation assay. The x axis represents the total number of days between the Delta and Omicron BA.1 dominance periods (eight days was the value used for generating all results in this study).

## Appendix 4. Neutralisation waning

**Figure 9.**
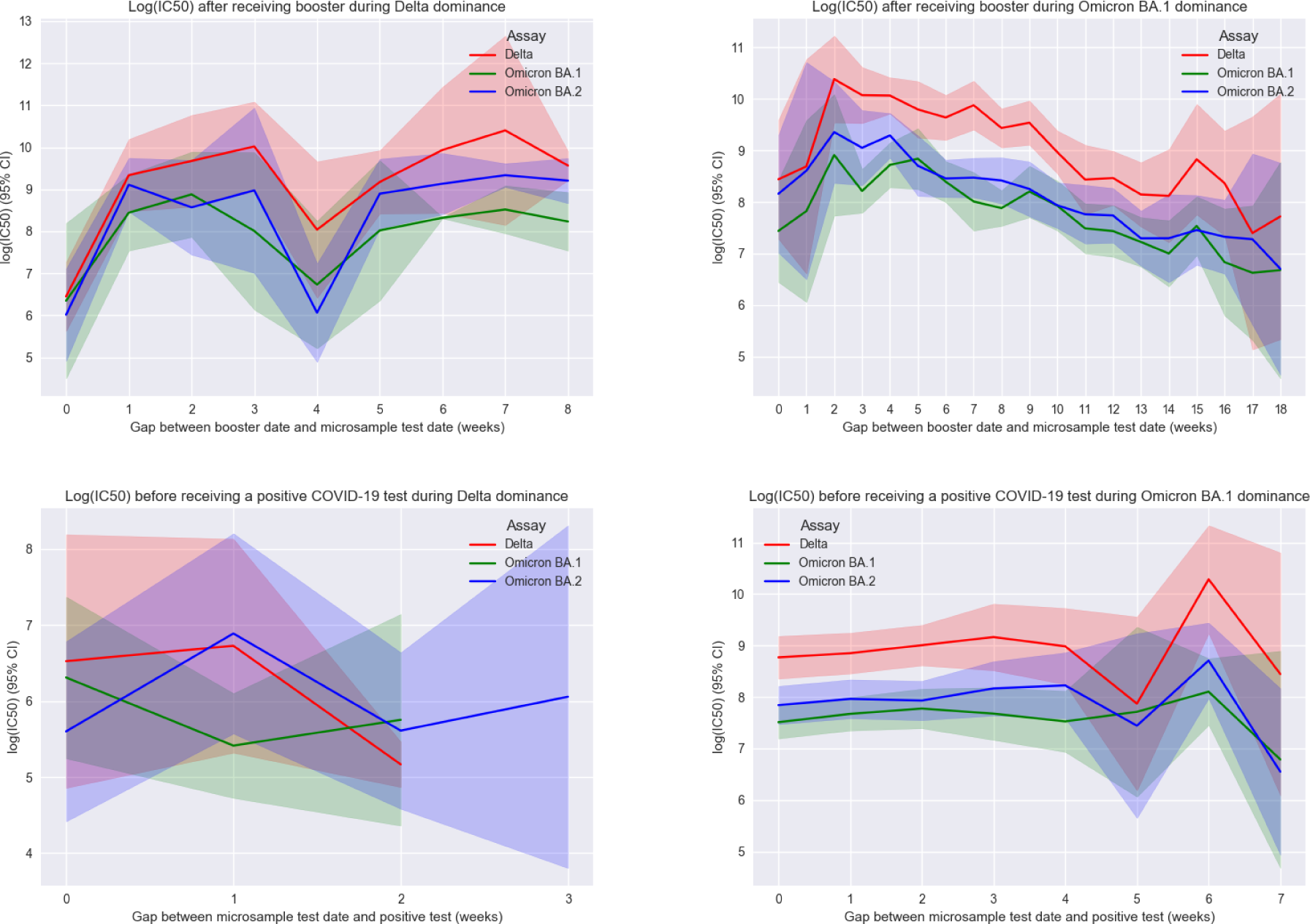
Neutralisation waning analysis

